# Constructing personalized characterizations of structural brain aberrations in patients with dementia and mild cognitive impairment using explainable artificial intelligence

**DOI:** 10.1101/2023.06.22.23291592

**Authors:** Esten H. Leonardsen, Karin Persson, Edvard Grødem, Nicola Dinsdale, Till Schellhorn, James M. Roe, Didac Vidal-Piñeiro, Øystein Sørensen, Tobias Kaufmann, Eric Westman, Andre Marquand, Geir Selbæk, Ole A. Andreassen, Thomas Wolfers, Lars T. Westlye, Yunpeng Wang, Alzheimer’s Disease Neuroimaging Initiative, Australian Imaging Biomarkers and Lifestyle flagship study of ageing

**Affiliations:** Department of Psychology, University of Oslo, Oslo, Norway; Norwegian Centre for Mental Disorders Research (NORMENT), Oslo University Hospital & Institute of Clinical Medicine, University of Oslo, Oslo,Norway; The Norwegian National Centre for Ageing and Health, Vestfold Hospital Trust, Norway; Department of Geriatric Medicine, Oslo University Hospital, Oslo, Norway; Computational Radiology & Artificial Intelligence (CRAI) Unit, Division of Radiology and Nuclear Medicine, Oslo University Hospital, Oslo, Norway; Oxford Machine Learning in NeuroImaging (OMNI) Lab, University of Oxford, UK; Institute of Clinical Medicine, University of Oslo, Oslo, Norway; Division of Radiology and Nuclear Medicine, Oslo University Hospital, Oslo, Norway; Department of Psychiatry and Psychotherapy, Tübingen Center for Mental Health, University of Tübingen, Germany; German Center for Mental Health (DZPG); Division of Clinical Geriatrics, Department of Neurobiology, Care Sciences, and Society, Karolinska Institutet, Stockholm, Sweden; Donders Institute for Brain, Cognition and Behaviour, Radboud University Medical Centre, Nijmegen, Netherlands; KG Jebsen Center for Neurodevelopmental Disorders, University of Oslo, Oslo, Norway

## Abstract

Deep learning approaches for clinical predictions based on magnetic resonance imaging data have shown great promise as a translational technology for diagnosis and prognosis in neurological disorders, but its clinical impact has been limited. This is partially attributed to the opaqueness of deep learning models, causing insufficient understanding of what underlies their decisions. To overcome this, we trained convolutional neural networks on structural brain scans to differentiate dementia patients from healthy controls, and applied layerwise relevance propagation to procure individual-level explanations of the model predictions. Through extensive validations we demonstrate that deviations recognized by the model corroborate existing knowledge of structural brain aberrations in dementia. By employing the explainable dementia classifier in a longitudinal dataset of patients with mild cognitive impairment, we show that the spatially rich explanations complement the model prediction when forecasting transition to dementia and help characterize the biological manifestation of disease in the individual brain. Overall, our work exemplifies the clinical potential of explainable artificial intelligence in precision medicine.

## Introduction

Since its invention in the 1970s, magnetic resonance imaging (MRI) has provided an opportunity to non-invasively examine the inside of the body. In neuroscience, images acquired with MRI scanners have been used to identify how the brains of patients with various neurological disorders differ from their healthy counterparts. Stereotypically, this has been done by collecting data from a group of patients with a given disorder and a comparable group of healthy controls, on which traditional statistical inference is applied to identify spatial locations of the brain where the groups differ ^1^. Typically, these locations are not atomic locations identified by spatial coordinates, but rather morphological regions defined by an atlas, derived from empirical or theoretical insights of how the brain is structured. Differences between groups are described using morphometric properties like thickness or volume of these prespecified regions. A major benefit of this approach is the innate interpretability of the results: on average, patients with a given disorder deviate in a specific region of the brain in a comprehensible manner. Furthermore, the high degree of localization offered by modern brain scans allows for accurate characterization of where and how the brain of an individual deviates from an expected, typically healthy, norm ^2^. However, the effects which are found are typically small ^3^ with limited predictive power at the individual level ^4,5^, which in turn has raised questions about whether these analytical methods are expressive enough to model complex mental or clinical phenomena ^6^. As an alternative, new conceptual approaches are proposed, advocating modelling frameworks with increased expressive power that allow for group differences through complex, non-linear interactions between multiple, potentially distant, parts of the brain ^7^, with a focus on prediction ^8^. Such modelling flexibility is naturally achieved with artificial neural networks (ANNs), a class of statistical learning methods that combines aspects of data at multiple levels of abstraction, to accurately solve a predictive task ^9^. However, while this often yields high predictive performance, e.g. by demonstrating clinically sufficient case-control classification accuracy for certain conditions, it comes at the cost of interpretation, as the models employ decision rules not trivially understandable by humans ^10^. When the goal of the analysis is clinical, supporting the diagnosis and treatment of someone affected by a potential disorder, this opaqueness presents a substantial limitation. Thus, development and empirical validation of new methods within clinical neuroimaging that combine predictive efficacy with individual-level interpretability is imperative, to facilitate trust in how the system is working, and to accurately describe inter-individual heterogeneity.

With more than 55 million individuals afflicted worldwide ^11^, over 25 million disability-adjusted life years lost ^12,13^ and a cost exceeding one trillion USD yearly ^14^, dementia is a prime example of a neurological disorders that incur a monumental global burden. Due to the global aging population the prevalence is expected to nearly triple by 2050 ^15^, inciting a demand for technological solutions to facilitate handling the upcoming surge of patients. Dementia is a complex and progressive clinical condition ^16^ with multiple causal determinants and moderators. Alzheimer’s disease (AD) is the most common form and accounts for 60%-80% of all cases ^11^. However, the brain pathologies underlying different subtypes of dementia are not disjoint, but often co-occur ^17–19^, and have neuropathological commonalities ^20^. The most prominent is neurodegeneration, occurring in both specific regions like the hippocampus, and globally across the brain ^21^, and inter-individual variations in the localization of atrophy has been associated with impairments in specific cognitive domains ^22,23^. Thus, the biological manifestation of dementia in the brain is heterogeneous ^24^, resulting in distinctive cognitive and functional deficits ^20^, highlighting the need for precise and personalized approaches to diagnosis. For patients with mild cognitive impairment (MCI), a potential clinical precursor to dementia, providing individualized characterizations of the underlying etiological disease at an early stage could widen the window for early interventions ^25^, alleviate uncertainty about the condition, and help with planning for the future ^26^.

In dementia, ANNs, and particularly convolutional neural networks (CNNs), have been applied to brain MRIs to differentiate patients from controls ^27,28^, prognosticate outcomes ^29^, and differentially diagnose subtypes ^30^. However, while research utilizing this technology has been influential, clinical translations are scarce ^31^. Where techniques for segmenting brain tumours or detecting lesions typically produce segmentation masks that are innately interpretable, predicting a complex diagnosis would entail compressing all information contained in a high-dimensional brain scan into a single number. Using deep learning, the decisions underlying this immense reduction are obfuscated, both from the developer of the system, the clinical personnel using it, and the patient ultimately impacted by the decision. This black box nature is broadly credited for the low levels of adoption in safety-critical domains like medicine ^32^. Responding to this limitation, explainable artificial intelligence (XAI) provides methodology to explain the behaviour of ANNs ^33^. The nature of these explanations varies, e.g. by what type of model is to be explained, what conceptual level the explanation is at, and who it is tailored for ^34,35^. In computer vision, XAI typically aims for post-hoc explanations of individual decisions, explaining why a model arrived at a given prediction for a given image. Explanations are often provided in a visual format, as a heatmap indicating how different regions of the image contribute to the prediction ^36^. Layerwise Relevance Propagation (LRP) is a variant of such a method, based on propagating relevance from the prediction-space, backwards through all layers of the model to the image-space, to form a relevance map ^37^. A major advantage of LRP is its intuitive interpretation: by construction, the total amount of relevance which denotes contribution to the prediction is kept fixed between layers. Thus, the relevance propagated back to an input voxel is directly indicative of the influence of that exact voxel to the prediction. Recently, several studies have applied both LRP and other explainable AI methods to dementia ^38^, finding that the heatmaps generally highlight regions known to change in dementia ^39–42^. However, the possibility of utilizing the fine-grained, individual, heatmaps produced by LRP to accurately characterize individualized disease manifestations has not been explored, despite its potential for supporting clinical decisions towards precision medicine ^41,43^.

In the present study, we applied techniques from deep learning and XAI on MRI scans of the brain to make explainable and clinically relevant predictions for dementia at the individual level (Figure 1). Using a state-of-the-art architecture for neuroimaging data, we trained CNNs to differentiate patients diagnosed with dementia from healthy controls based on T1-weighted structural MRIs. We implemented LRP on top of the trained models to form a computational pipeline producing individual-level explanations in the form of relevance maps alongside the model predictions. The relevance maps were validated in a subset of dementia patients, both in a qualitative comparison with existing knowledge of the anatomical distribution of structural aberrations, and in a quantitative, predictive context. Next, we applied the pipeline to a large, longitudinal dataset of MCI patients to create individual morphological records, a proposed data format for tracking and visualizing disease progression. Finally, we investigated the clinical utility of these records for stratifying patients, both in terms of their specific clinical profile, and progression of the disease. To facilitate reproducibility and improve the translational value of our work, the trained models and the complete explainable pipeline is made accessible on GitHub.

**Figure 1:**
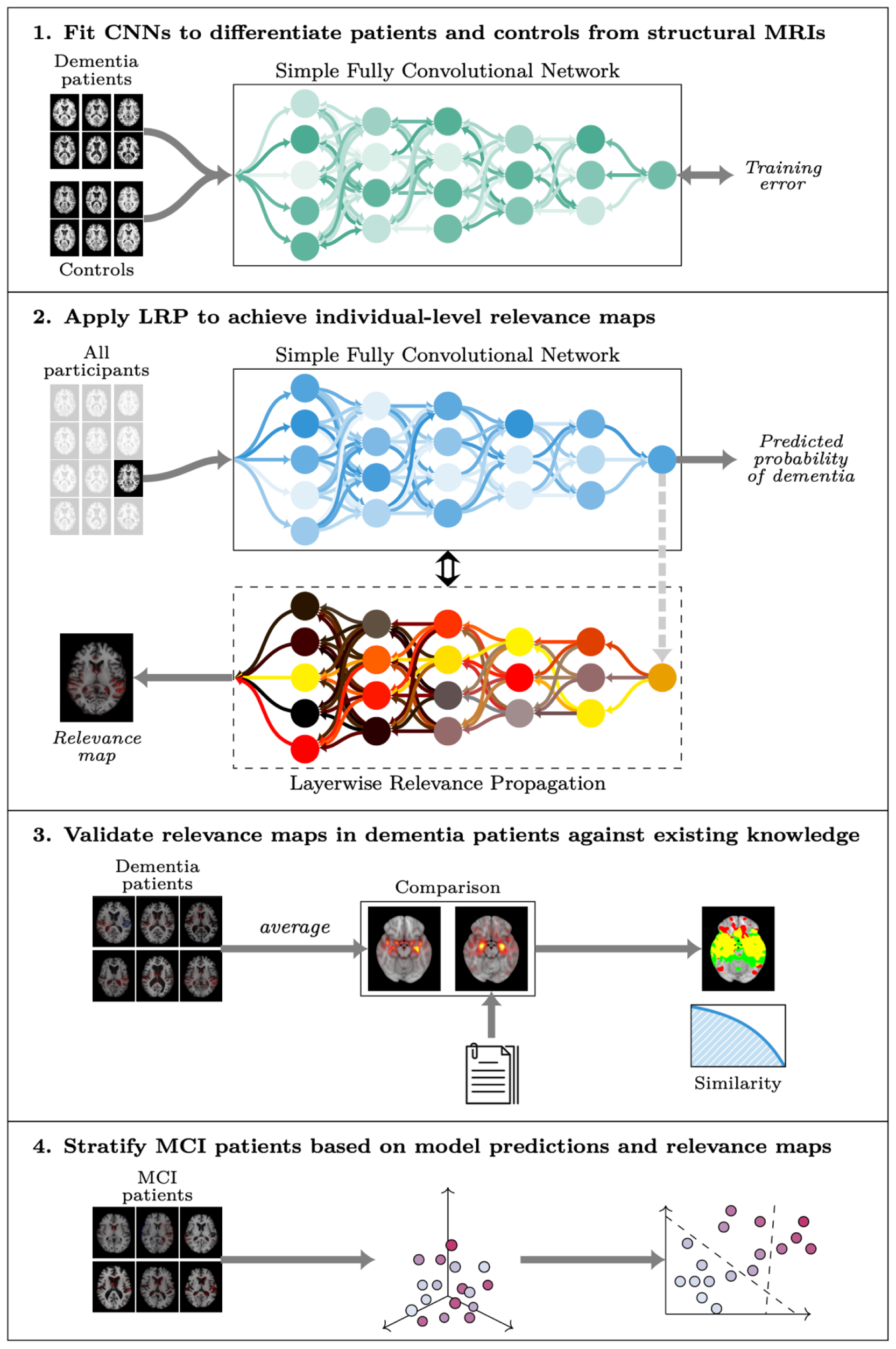
Overview of the modelling process. The modelling process consisted of four sequential steps. First, we fit multiple Simple Fully Convolutional Networks to classify dementia patients and healthy controls based on structural MRIs. Then we applied the best models to generate out-of-sample predictions and relevance maps for all participants. Next, we validated the relevance maps against existing knowledge using a meta-analysis to generate a statistical reference map. Finally, we employed the full pipeline in an exploratory analysis to stratify patients with mild cognitive impairment (MCI).

## Results

We compiled MRI data from multiple sources (Supplementary Table 1) into a dataset of heterogeneous dementia patients (n=854, age range=47-95, 47% females, Table 1) based on various diagnoses (Probable AD, vascular dementia, other/unspecified dementia) and diagnostic criteria for inclusion (Supplementary Table 2), and a set of controls strictly matched on site, age, and sex of equal size. We trained multiple CNNs to differentiate between the groups, employing a cross-validation approach utilizing all available timepoints for participants in three training folds and a single randomly selected timepoint for participants in separate validation and test folds. When stacking the out-of-sample predictions for all participants from all folds together (n=1708), for each fold using the model with the best validation performance, we observed satisfactory discrimination with a combined area under the receiver operating characteristics curve (AUC) of 0.908 (0.904-0.920 split across folds, Supplementary Figure 1), and an accuracy of 84.95% (83.04%-87.13%, Supplementary Table 3). This is slightly below with what is commonly achieved in similar studies classifying a specific subtype (typically AD) in a single dataset ^28^.

**Table 1:** Cohorts. Key characteristics of the cohorts used for training and testing the models, and further exploratory analyses.

| CNN training and cross-validation |  |  |  |
| --- | --- | --- | --- |
| Cohort | Participants | Mean age (±std) | Sex (F/M) |
| Healthy controls | 854 | 75.13±7.81 | 401/453 |
| Dementia patients | 854 | 74.82±7.84 | 401/453 |
| **Total** | **1708** | **74.98±7.82** | **802/906** |
| Downstream prognostic and correlational analyses |  |  |  |
| Improved MCI | 80 | 71.18±8.14 | 37/43 |
| Stable MCI | 754 | 74.63±**7.66** | 324/430 |
| Progressive MCI | 304 | 75.60±**7.46** | 124/180 |
| **Total** | **1138** | **74.67±7.73** | **485/653** |

### Relevance maps highlight predictive brain regions in individuals with dementia

Based on the classifiers with the highest AUCs in the validation sets, we built an explainable pipeline for dementia prediction,*LRP_dementia_*, using composite LRP^44^, and a strategy to prioritize regions of the brain that contributed positively towards a prediction of dementia in the explanations. Using this pipeline, we computed out-of-sample relevance maps for all participants by applying the model for which the participant was unseen. Qualitatively, these maps corroborated known anatomical locations with structural aberrations in dementia, while still allowing for inter-individual variation (Supplementary Figure 2). We confirmed this apparent corroboration quantitatively by comparing a voxel-wise average map 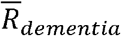 (Supplementary Figure 3), containing positive relevance from all correctly predicted dementia patients, with a statistical reference map *G* (Supplementary Figure 4) from an activation likelihood estimation (ALE) meta-analysis ^45^, methodology established by an earlier study ^40^. For sanity checks, we also computed average maps from three alternative pipelines, 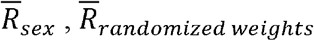 and 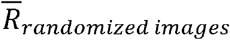. The comparisons with the reference map were done by binarizing the maps on both sides of the comparison at various thresholds and measuring the Dice overlap (Figure 2a). For the three alternative pipelines the amount of overlap decreased monotonically as the binarization threshold rose (Figure 2b), whereas for 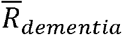 it stabilized as the maps grew sparser, indicating its higher similarity with *G*. This effect was reaffirmed by a normalized cross-correlation ^46^ of 0.64 for 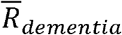, compared to 0.41, 0.40 and 0.12 of 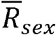, 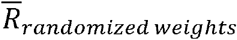 and 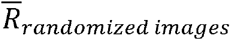 respectively. In addition, we performed a region-wise, qualitative comparison of 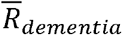 and, also yielding general agreement (Figure 2c), with the most important regions in both maps being the nucleus accumbens, the amygdala, and the parahippocampal gyrus. Next, we tested the importance of the detected regions in a predictive context, by applying an iterative mask-and-predict procedure. For each participant, we produced a baseline dementia-prediction 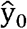 and relevance map *R_task_* for each pipeline *LRP_task_*. We then iteratively masked out the most important regions of the image according to the relevance map and recorded how the prediction changed as a function of the occlusion (Figure 2d). Using only true positives, the predictions should ideally start out at approximately 1.0 (empirically found to be 0.89 on average) and trend towards 0.5 (random prediction) as a larger proportion of the image is occluded. The rate of decline is indicative of whether the masked regions contain information essential for the classifier to classify the image correctly. Over 20 integration we observed that the predictions based on maps from both *LRP_dementia_,LRP_sex_* and *LRP_randomized weights_* decreased, but *LRP_dementia_* at a distinctly steeper rate than the rest (Figure 2d). To quantify this observation we calculated an area over the perturbation curve (AOPC) of 0.231, 0.009, -0.001 and 0.002 for *LRP_dementia_, LRP_sex,_ LRP_randomized images,_ LRP_randomized weights_* respectively. Taken together, these results demonstrate that our pipeline generates maps with relevance in brain regions associated with changes in dementia.

**Figure 2:**
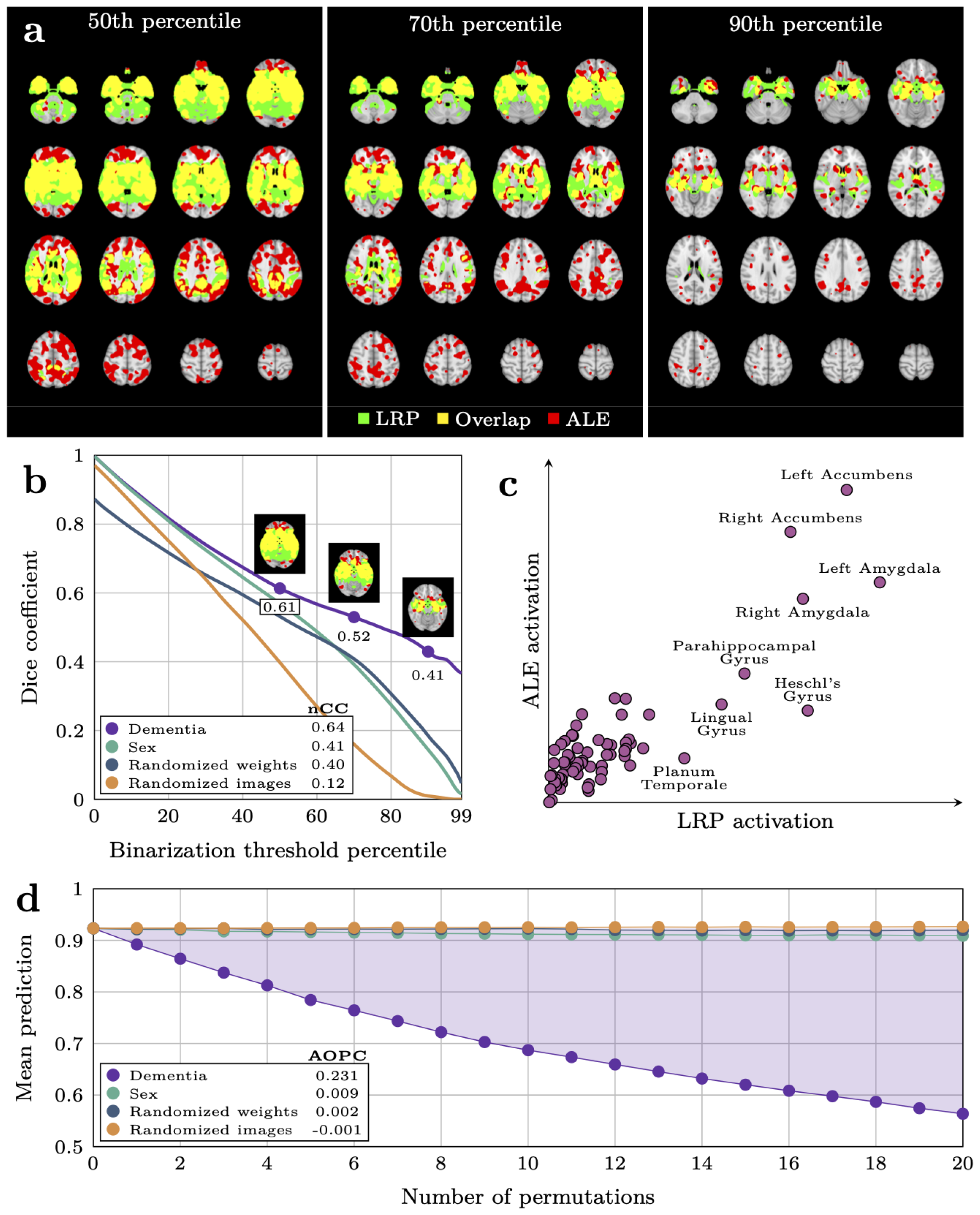
Validation of relevance maps from the dementia pipeline compared with three alternative pipelines. **a** Visualization of the comparison between the binarized average relevance 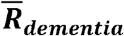 map _dementia_ from the dementia-pipeline and the binarized statistical reference map **G** from GingerALE, at different thresholds for binarization. **b** Overlap between the four average relevance maps 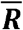 from our four pipelines and as a function of the binarization threshold. The numbers in the legend denote the normalized Cross Correlation (nCC) for each pipeline **c** Mean voxel-wise activation in 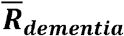and **G**, grouped by brain region. **d** Average participant-wise prediction from the dementia model after iteratively masking out regions of the image according to relevance maps from the four pipelines. Area over the permutation curve (AOPC) for the dementia map is indicated by the shaded area and denoted in the legend for all pipelines.

### Output from the explainable dementia pipeline has prognostic value for MCI patients

For the MCI patients (n=1256, timepoints=6448), previously unseen by all models, we built an averaging ensemble to procure a singular out-of-sample prediction and relevance map per patient per timepoint. Put together, we let this represent a morphological record (illustrated in Figure 4) visualizing the absolute quantity (indicated by the prediction) and location (indicated by the relevance map) of dementia-related pathology detected by the models over time. Qualitatively, both predictions and maps were relatively stable within a participant over time, while allowing enough variation to compose what resembled a trajectory. To investigate the prognostic value of our proposed morphological records we divided the MCI patients into three subgroups based on their trajectories in the follow-up period: those who saw improvement of their condition (n=80), those with a stable diagnosis throughout (sMCI, n=754), and those who progressed into dementia (pMCI, n=304). The remaining (n=118) had either a non-MCI diagnosis at the first timepoint, or a more complex diagnostic trajectory (e.g MCI -> AD -> CN) and were excluded from subsequent analyses. We observed that the predictions in the first group were generally very low (mean 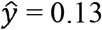, Supplementary Figure 5a), indicating that the models detected little, if any, evidence of dementia in these participants. For the stable patients the mean prediction was higher (mean 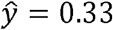), but still below the classification threshold of 0.5, whereas in the progressive group the model predicted the average patient to already have dementia (mean 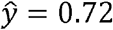). Importantly, this was also true when considering only timepoints before these patients received the clinical diagnosis (mean 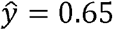, Supplementary Figure 5b), suggesting that the model found evidence of the disorder before the clinical symptoms surpassed the diagnostic threshold. To formally delineate the differences in predictions leading up to the potential diagnosis, we combined the improving and stable patients into a non-progressive group (nMCI, n=834), and sampled patients to match the progressive group based on their visiting histories, leading up to a terminal diagnosis timepoint (or a constructed non-diagnosis timepoint in the non-progressive group). In this matched dataset (n=550) we applied a linear mixed model controlling for age and sex and observed that the group difference was even greater than what we previously observed (β = 0.47, p = 6.05 × 10^−71^, Figure 3a, Supplementary Table 4). Furthermore, we observed a significant difference in longitudinal slopes (β = 0.05 increase in prediction per year, p = 8.14 × 10^−17^) indicating a greater rate of brain change detected by the model in those who would be diagnosed with dementia at a later point in time.

**Figure 3:**
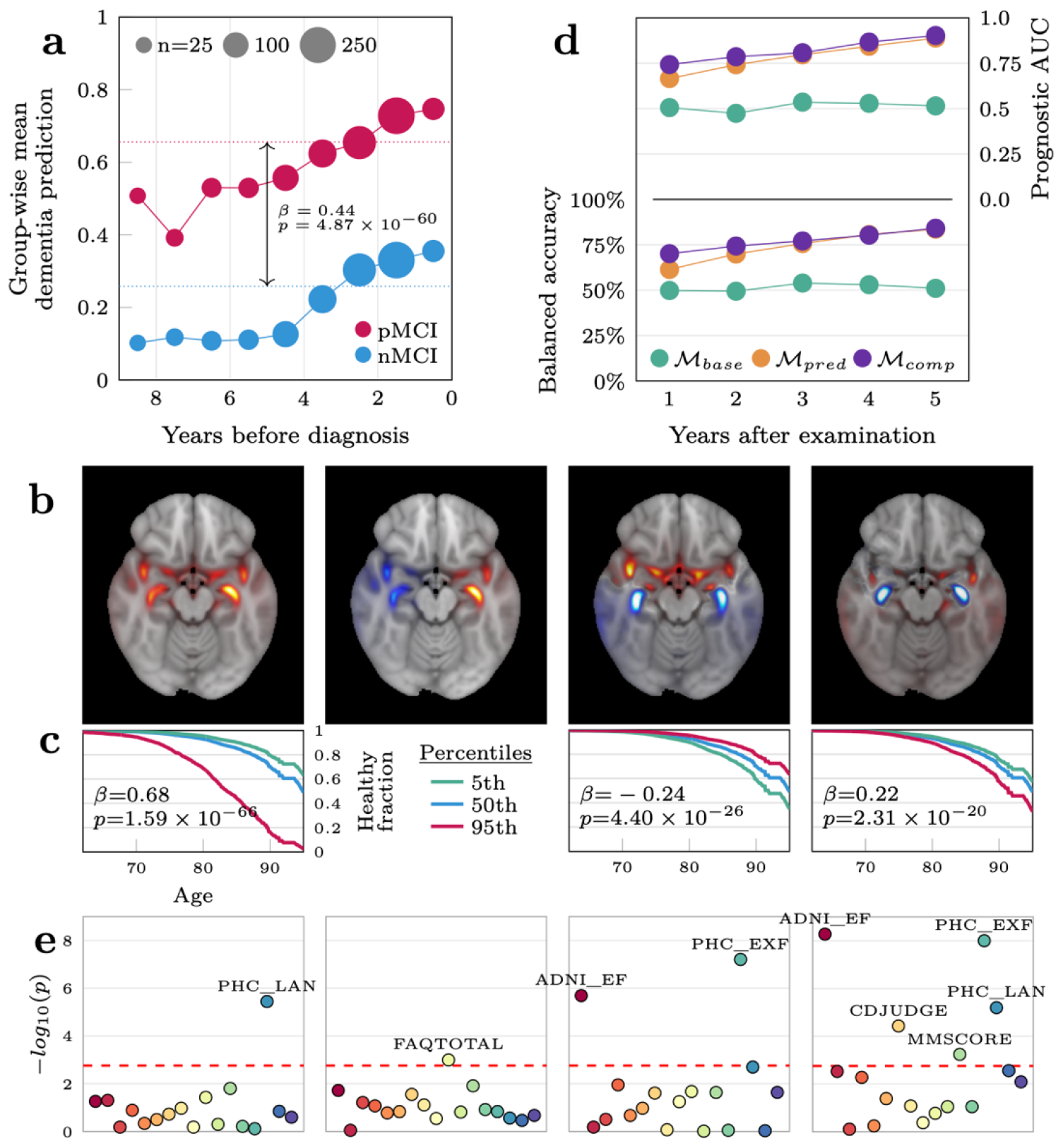
Utility of the dementia pipeline for predicting progression and characterizing individual-level deviations in the mild cognitive impairment cohort. **a** Group-wise mean predictions from the dementia-model in the progressive and non-progressive groups in the years before a diagnosis was given. **b** The four first voxel-wise components of the principal component analysis plotted in MNI152-space. **c** Survival curves for the average MCI patient (blue) and fictitious patients at the extreme percentiles of the span for each component. The second component was not significant and is not shown. **d** Predictive performance of the three models predicting progression in the years following the MRI examination. The baseline model () included only sex and age as covariates, the next model included the prediction from the dementia classifier as a predictor, while the final model also added the component vectors representing the relevance maps. **e** Significance levels of correlations between the each of the four PCA components and various cognitive measures. The six annotated measures are composite language (PHC_LAN) and executive function (PHC_EXF) scores from the ADSP Phenotype Harmonization Consortium, total score from the Functional Activities Questionnaire (FAQTOTAL), composite executive function score from UW – Neuropsych Summary Scores (ADNI_EF), clinical evaluation of impairment related to judgement and problem solving (CDJUDGE) from the Clinical Dementia Rating, and an overall measure of cognition from the Mini-Mental State Examination (MMSCORE, commonly referred to as MMSE).

**Figure 4:**
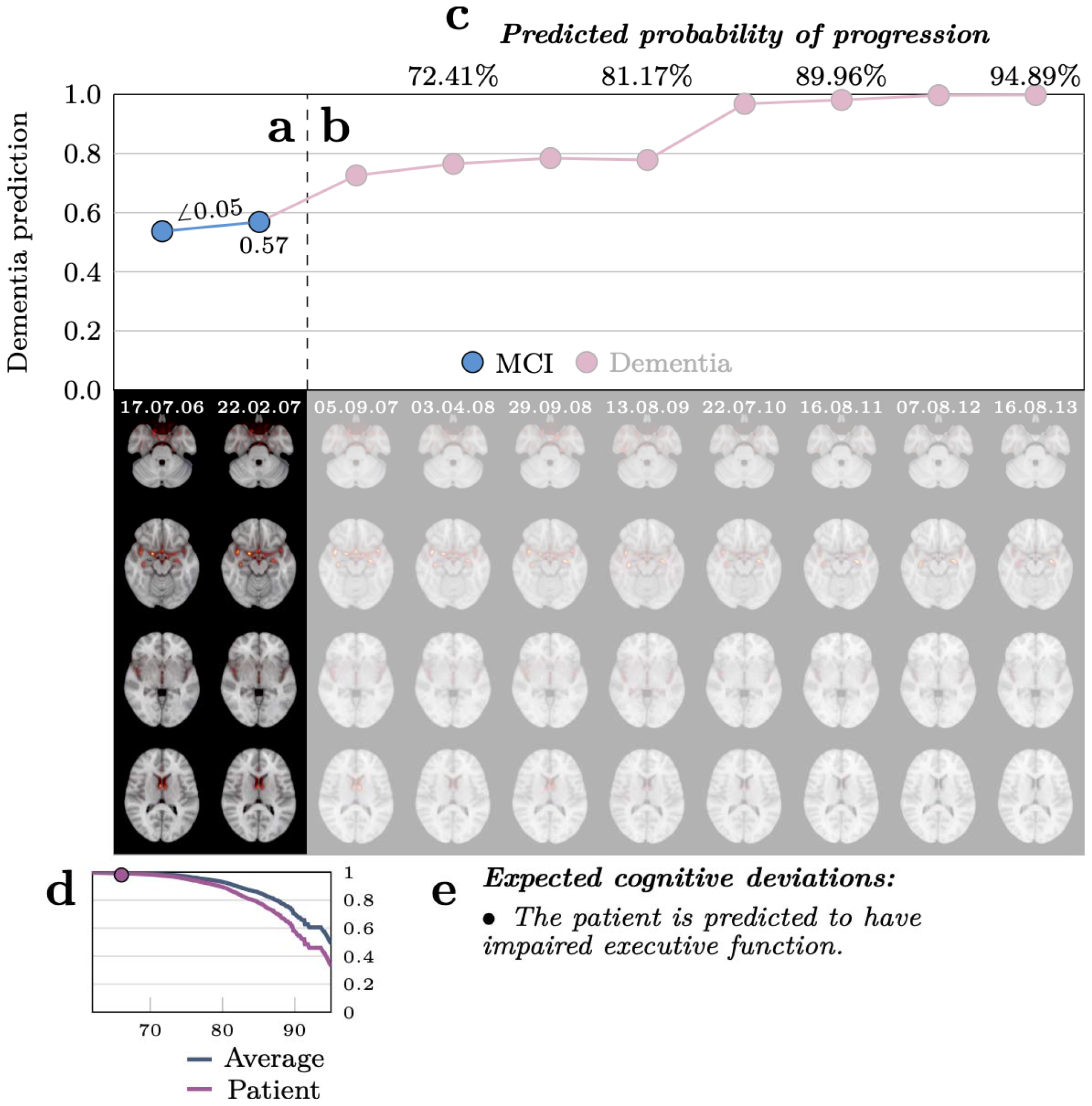
A visualization of the proposed morphological record for a randomly selected progressive MCI patient that was held out of all models and analyses. **a** The top half shows the prediction from the dementia model at each visit, while the bottom part displays the relevance map underlying the prediction. The opaque sections (including **c, d,** and **e**) contain information accessible at the imagined current timepoint (22.02.07) to support a clinician in a diagnostic procedure. The angle () represents the change in dementia prediction per year based on the first two visits. **b** Translucent regions reveal the morphological record for the remaining follow ups in the dataset, thus depicting the future. The ground truth diagnostic trajectory is encoded by the colour of the markers. **c** Predicted probabilities of progression at future follow-ups based on the prediction and relevance map at the current timepoint. **d** Survival curve of the patient compared to the average MCI patient calculated from the prediction and relevance map. The marker indicates the location of the patient at the current timepoint. **e** A list of cognitive domains where the patient is predicted to significantly differ from the average based on the prediction and relevance map.

The large group differences in the dementia predictions leading up to a potential diagnosis suggests this as a biomarker with innate prognostic value, yet the most salient part of our morphological records were the relevance maps. Thus, we performed exploratory analyses based on these to further differentiate the non-progressive and progressive groups and characterize both inter- and intra-group heterogeneity. However, given the high dimensionality of the maps and the relatively small number of patients, we first applied a principal component analysis (PCA) to relevance maps from all MCI patients, effectively compressing their information content into a smaller set of characteristic variables encoding facets of the maps, enabling the subsequent analyses. We retained the 64 components that explained the largest amount of variance and observed that they qualitatively clustered into three overarching categories. The first component was a generic component detecting general presence of relevance, resembling the average map from dementia patients, and thus made up a cluster by itself. The next cluster was comprised of the subsequent three components that captured high level, abstract patterns of relevance, namely differences in lateralization, along the sagittal axis and in subcortical regions (Figure 3b). The final cluster consisted of the remaining 60 components that captured specific, intricate patterns of presence/non-presence of relevance in regions revealed in the preceding analyses (Supplementary Figure 6). To investigate the potential of using the relevance maps for prognosis, we first performed a survival analysis using a Cox proportional hazards model where getting a diagnosis was considered the terminal event.

Specifically, we modelled the fraction of the population without a diagnosis as a function of age and used the subject-wise loadings of *c*_t_ as predictors. After Benjamini-Hochberg correction, 37 of these components were significantly associated with staying undiagnosed (Figure 3c and Supplementary Table 5). However, we observed a correlation between the singular dementia prediction 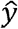 and the absolute magnitudes of these components (Supplementary Figure 7), indicating that the associations in the survival analysis could be induced by differences in the prediction rather than variability in the relevance maps. To mitigate this concern, we fit an equivalent model while stratifying on 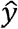, observing that 29 associations remained significant, and that all coefficients had the same sign. Nonetheless, this analysis did not account for the predictions and relevance maps changing within a participant over time, so we reframed the question in a purely predictive setting, constructed to bear resemblance to a clinical scenario,using the same participants (nMCI=834, pMCI=304, total n=1138). For each MCI patient *p* at each timepoint *t* we asked whether we were able to predict, at yearly intervals γ up to five years into the future, whether *p* had progressed into dementia, using information from *LRP_dementia_* available at *t*. Importantly, all timepoints for all these participants were unseen by the dementia-model, yielding out of sample predictions and relevance maps from *LRP_dementia_*, and we employed nested cross-validation to ensure the progression predictions were also out-of-sample. First, we fit a baseline model ℳ*_base_* with age and sex as predictors, showing no predictive efficacy at any timepoint (all AUCs ≈ 0.5, Supplementary Table 6), indicating that the dataset was not biased with respect to these variables. When adding the prediction from the dementia model 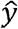*_t_* as a predictor in model ℳ*_pred_* we saw large improvements in prognostic efficacy at all yearly intervals, culminating with a fold-wise mean AUC of 0.889 after five years (Figure 3d). In the final model, ℳ*c_omp_*, also including the component vector *c_t_* as predictors, we saw further improvements for all years, peaking at 0.903 after five years (p = 0.035 when compared to _ℳ*pred*_ in a Wilcoxon signed-rank test across the outer folds). Overall, our best performing model predicted progression to dementia after five years with an AUC of 0.903, an accuracy of 84.1%, a positive predicted value of 0.92, a sensitivity of 0.82 and a specificity of 0.86 (Table 2).

**Table 2:** Predictive performance of the three models predicting progression five years into the future. The baseline model ℳ_base_ used only age and sex as covariates. ℳ_pred_ also added the prediction from the dementia model at the current timepoint as a predictor, while ℳc_omp_ additionally included the component vector c_t_ encoding information from the relevance maps.

| Model | AUC | Balanced accuracy | PPV | Sensitivity | Specificity |
| --- | --- | --- | --- | --- | --- |
| $\mathcal{M}_{base}$ | 0.515 | 51.05% | 0.14 | 0.09 | 0.93 |
| $\mathcal{M}_{pred}$ | 0.889 | 83.61% | 0.91 | 0.83 | 0.84 |
| $\mathcal{M}_{comp}$ | 0.903 | 84.1% | 0.92 | 0.82 | 0.86 |

### Facets of the relevance maps are associated with cognitive impairments in distinct domains

Finally, we tested whether common features found in the relevance maps, represented by the PCA component, were correlated with impairments in distinct cognitive and functional domains. We extracted 17 summary measures from 7 neuropsychological tests (Supplementary Table 7 and 8), performed approximately at the same time as an MRI examination, and tested for associations with the subject-wise loadings of *c_t_* in 733 MCI patients using linear models. After FDR correction, while correcting for age, sex and 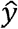, we found 48 significant correlations between 18 unique components and 14 of the cognitive measures (Figure 3e). Component 30 and the aggregate score from the Functional Activities Questionnaire (FAQTOTAL) had the highest number of significant hits among the components and measures respectively, both with six passing the threshold. Most importantly, the components showed distinct patterns of associations with the different cognitive measures. To ensure the significant associations were not driven by collinearity between components c*_i_* and 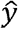, we ran an equivalent analysis without including 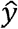 as a predictor, observing that only 5/48 of the previously significant hits had coefficients with the opposite sign. To summarize, the spatial features captured in our relevance maps, and subsequently in our component vectors, were associated with distinct patterns of performance on neuropsychological tests relevant for characterizing phenotypic heterogeneity in dementia patients (Supplementary Figure 8).

## Discussion

Given the huge burden of disease and expected increase in prevalence, innovative technological solutions for clinical decision making in dementia diagnostics and prognostics is urgently needed. Although commonly referred to as a homogenous condition or split into a few subtypes based on aetiology or pathophysiology ^17^, dementia patients exhibit unique and complex deficiencies, disease trajectories, and cognitive deficits. To explore the potential of brain MRI and XAI to characterize heterogeneity in the brain underpinnings of dementia, we trained neural networks to differentiate dementia patients from healthy individuals, and derived relevance maps using Layerwise Relevance Propagation to explain the individual-level decisions of the classifier. The relevance maps were specific to the individual, spanned regions that were predictive of dementia and corroborated existing knowledge of the anatomical distribution of structural aberrations. In a cohort of MCI patients, it enabled characterization and differentiation of individual-level disease manifestations and trajectories linked to cognitive performance in multiple domains. While further validations in clinical contexts are needed, our XAI pipeline for dementia demonstrates how advanced predictive technology can be employed by clinicians to monitor and characterize disease development for individual patients.

There is a multitude of XAI techniques available for explaining the decisions of an image classifier, many of which have yielded promising results for dementia classification ^38^. We employed LRP due to its straightforward interpretation as well as earlier studies indicating robustness ^47^ and specificity ^42^, properties we consider integral in a clinical decision support system. But while procuring explanations that are *ipso facto* meaningful is an important step towards adoption of AI in clinical neuroimaging, it is not in itself sufficient. There is a host of predictive models that are trivially explainable, but not understandable ^48^, and there is genuine concern that XAI will lead to another level of systems that are formally well-defined, but opaque and obscure, and thus practically useless ^49^. Thus, empirical explorations are imperative to investigate the nature of these explanations, examine how they may be useful and build essential trust ^50^. In our validation, we observed that the explanatory maps produced by the dementia pipeline were more predictive and showed distinctly more agreement with existing knowledge of pathology than those produced by the three alternative pipelines. Given limitations that have been exposed in such methods earlier ^51,52^ these validations are crucial, and observing that our results both corroborate earlier evidence ^40^ and extend upon it, provides confidence that the explanations derived from the model are meaningful. However, we emphasize that the ultimate validation should happen in actual implementations of the technology in end-user systems, with clinical personnel applying it in clinical scenarios on realistic data.

We continued beyond validating the relevance maps by proposing them as a potential epistemic and clinical tool to characterize individual facets of dementia. To this end, we explored if the maps contributed to predicting imminent progression from MCI to dementia, and correlated them with different cognitive measures, extending upon the current literature ^38^. In both analyses we found evidence, although modest, that the maps are informative beyond the predictions of the model. To illustrate the potential of the pipeline for clinical decision making we compiled its output into a proposed morphological record (visualized for a single patient in Figure 4) that can help clinicians localize morphological abnormalities during a diagnostic process. Identifying subtle pathophysiology through deep phenotyping could have a huge potential for charting the heterogeneity of dementia, providing precise biological targets to guide future research. Furthermore, for the individual patient, it can support personalized diagnosis to identify appropriate disease-modifying treatments, and in the future, hopefully, accurate therapeutic interventions.

The regions with the highest density of relevance in our maps were the nucleus accumbens, amygdala and the parahippocampal gyrus, all of which are strongly affected in dementia ^53–55^. While the two latter corroborate the established involvement of the medial temporal lobe ^56^ it is surprising that the hippocampus does not appear in our analyses, as it has frequently in similar studies ^38,41,42^. While this could be caused by actual localization of pathology ^57^ we consider it more likely to be related to the internal machinery of the model. Specifically, the CNN relies on spatial context to identify brain regions before assessing their integrity, utilizing filters that span areas of the image larger than those containing the region itself. In the backwards pass, LRP uses these filters, and thus the localization of relevance is not necessarily voxel precise. Furthermore, we believe the model broadly can be seen as an atrophy detector, which necessarily entails looking for gaps surrounding regions instead of directly at the regions themselves. Therefore, while the relevance maps provide important information, they depend on contextual information and thus rely on interpretation from clinicians to maximize their utility in clinical practice.

We focused our analyses mainly on the relevance maps, but the results with largest, immediate, potential for clinical utility were the predictions from the dementia classifier. Other studies have shown the efficacy of machine learning models in differentiating dementia patients and healthy controls ^28^, but it is intriguing that we see a large discrepancy in the predictions of the progressive and non-progressive MCI patients many years before the dementia diagnosis is given. This corroborates findings from theory-driven studies ^58^ and a recent deep learning study ^27^, implying detectable structural brain changes many years before the clinical diagnosis is given. This gives hope for advanced technology to contribute to early detection and diagnosis through MRI based risk scores, in our case supported by a visual explanation. If curative treatments prove efficacious and become accessible, early identification of eligible patients could be imperative ^59^. Furthermore, timely access to interventions have shown efficiency in slowing the progress of cognitive decline ^60^, in addition to improving the quality of life for those afflicted and their caregivers ^26,61^. Widely accessible technology that allows for early detection with high precision could play a key role in the collective response to the impending surge of patients and provide an early window of opportunity for more effective treatments.

While our results show a great potential for explainable AI, and particularly LRP, as a translational technology to detect and characterize dementia, there are limitations to our study. First, there are technical caveats to be aware of. Most importantly, there is an absolute dependence between the predictions of our model and the relevance maps. In our case, when we qualitatively assessed the relevance maps of the false negatives, they were indistinguishable from the true negatives. This emphasizes the fact that when the model is wrong, this is not evident from the explanations. Next, while the maps contain information sufficient to explain the prediction, they are not necessarily complete. Thus, they don’t contain all evidence in the MRI pointing towards a diagnosis, a property which could prove essential for personalization. We have addressed this problem through pragmatic solutions, namely ensembling and targeted augmentations, but theoretical development of the core methodology might be necessary to theoretically guarantee complete maps. Beyond the fundamental aspects of LRP, there are weaknesses to the present study that should be acknowledged. First, the dataset with dementia patients portrayed as heterogeneous mostly consists of ADNI and OASIS data, and thus patients with a probable AD diagnosis (although clinically determined). Thus, while we consider it likely, it is not necessarily true that the dimension of variability spanning from healthy controls to dementia patients portrayed by our model has the expressive power to extrapolate to other aetiologies. To overcome this in actual clinical implementations, we encourage the use of datasets that are organically collected from subsets of the population that are experiencing early cognitive impairments, for instance from memory clinics. Furthermore, it is not trivial to determine whether a clinical, broad, dementia-label is an ideal predictive target for models in clinical scenarios. Both ADNI and AIBL contain rich biomarker information with multiple variables known to be associated with dementia, such as amyloid positivity. It would be intriguing to see studies methodologically similar to ours with a biological predictive target, and we encourage investigations into whether this supports and complements the results we have observed here. Another limitation with the present study is out-of-sample generalization, especially related to scanners and acquisition protocols. Although we utilize data from many sites, which we have earlier shown to somewhat address this problem ^62^, in combination with transfer learning, we did not explicitly test this by e.g., leaving sites out for validation. Again, we advise that clinical implementations should be based on realistic data, and thus at least be finetuned towards data coming from the relevant site, scanner, and protocol implemented in the clinic ^63^. This also includes training models with class frequencies matching those observed in clinical settings, instead of naively balancing classes as we have done here. Next, we want to explicitly mention the cyclicality of our mask-and-predict validation. In a sense it trivially follows that regions that are considered important by a model are also the ones that are driving the predictions, and thus it is no surprise that the relevance maps coming from the dementia model are more important to the dementia model than the maps coming from e.g., the sex model. We addressed this by alternating the models for test and validation, but fully avoiding this circularity would require disjunct datasets, and more and larger cohorts. Finally, we highlight the potential drawbacks of including the improving MCI patients alongside the stable in the progression models. We believe this accurately depicts a realistic clinical scenario, where diagnostic and prognostic procedures happen based on currently available clinical information.

However, that these patients improve could indicate that their condition is not caused by stable biological aberrations. This could oversimplify the subsequent predictive task, inflating our performance measures. In summary, the predictive value we observed for the individual patient must be interpreted with caution. However, our extensive validation approach as well as our thorough explanation of the method and its limitations, and training on large datasets, provide a first step towards making explainable AI relevant for clinical decision support in neurological disorders. Nonetheless, it also reveals a complicated balance between validating against existing knowledge and allowing for new discoveries. In our case, confirming whether small details revealed in the relevance maps are important aspects of individualization or simply intra-individual noise requires datasets with a label-resolution beyond what currently exists. Thus, we reiterate our belief that the continuation of our work should happen at the intersection between clinical practice and research ^64^, by continuously collecting and labelling data to develop and validate technology in realistic settings.

To conclude, while there are still challenges to overcome, our study provides an empirical foundation and a roadmap for implementations of brain MRI based explainable AI in personalized clinical decision support systems. Specifically, we show that deep neural networks trained on a heterogenous set of brain MRI scans can predict dementia, and that their predictions can be made human interpretable. Furthermore, our pipeline allows us to reason about structural brain aberrations in individuals showing early signs of cognitive impairment by providing personalized characterizations which can subsequently be used for precise phenotyping and prognosis, thus fulfilling a realistic clinical purpose.

## Materials and Methods

### Data

All data used in the present study have been obtained from previously published studies which have been approved by their respective institutional review board or relevant research ethics committee.

To train the dementia models we compiled a case-control dataset from seven different sources (Supplementary Table 1), consisting of patients with a dementia diagnosis and healthy controls from the same scanning sites. Because of the different diagnostic criteria used in the original datasets we applied different rules to achieve a singular, heterogeneous dementia label (Supplementary Table 2). We extracted all participants with a dementia-diagnosis at all timepoints to comprise the patient group (n=854). Then, for each unique proxy site (In ADNI, due to the large number of scanners and acquisition protocols, and the work put into unifying them, we used field strength as a proxy for site), sex, and age-bin spanning 10 years, we sampled an equal number of healthy controls to form the matched control set (total n=1708, Table 1). Lastly, before modelling, we split the data into five equally sized folds stratified on diagnosis, site, sex, and age, such that all timepoints for a single participant resided in the same fold.

For the MCI dataset we started with all participants from all ADNI waves with an MCI diagnosis (subjective memory complaint, MMSE between 24 and 30, CDR>0.5 with memory box>0.5, Weschler Memory Scale-Revised <9 for 16 years of education, <5 for 8-15 years of education and <3 for 0-7 years of education) ^65^, on at least one timepoint. These were 12661 images from 6448 visits for 1256 participants, none of which were used for model training. This selection criterion ensured all participants had an MCI diagnosis at one point in time, though it did not limit us to only those timepoints. Thus, in addition to those with a consistent, stable, MCI diagnosis (sMCI), we had a variety of diagnostic trajectories, including those transitioning from normal cognition to MCI, MCI to AD (pMCI) and various other combinations. Before the subsequent analyses we discarded all participants without an MCI diagnosis initially, and everyone with ambiguous trajectories (e.g. MCI->CN->AD), leaving 5607 visits from 1138 participants.

From these two datasets we extracted T1-weighted structural MRI data for each participant at each timepoint to use as inputs for the subsequent predictive models. Prior to modelling, the raw images were minimally processed using a previously developed pipeline ^2^/21/2024 12:12:00 PM relying on FreeSurfer v5.3 and FSL v6.0 ^66^ to perform skullstripping ^67^ and linear registration to MNI152-space ^68^ with six degrees of freedom. Consequently, the processed images consisted of normalized voxel values from the raw images, registered to a common spatial template and contained minimal non-brain tissue.

### Modelling

All dementia models were variants of the PAC2019-winning simple fully convolutional network (SFCN) architecture ^69,70^, modified to have a single output neuron with a sigmoid activation. The architecture is a simple, VGG-like convolutional neural network with 6 convolutional blocks and approximately 3 million parameters. We initialized the model with weights from a publicly accessible brain age model previously shown to have superior generalization capabilities when dealing with unseen scanning sites and protocols ^62^. The models were trained on a single Nvidia A100 GPU with 40GB of memory, Tensorflow 2.6 ^71^ through the Keras interface ^72^. We used a vanilla stochastic gradient descent (SGD) optimizer with a learning rate defined by the hyperparameter settings (see next section), optimizing the binary cross-entropy loss. All models ran for 160 epochs with a batch size of 6, and for each run the epoch with the lowest validation loss was chosen. Varying slightly depending on the hyperparameters, a single model trained in approximately 4 hours.

For each hold-out test fold we trained models on three of the remaining folds and validated on the fourth, akin to a cross-validation with an additional out-of-sample test set, to achieve out-of sample predictions for all 1708 participants while allowing for hyperparameter tuning. The hyperparameters we optimized were dropout *d* ∈{0.25, 0.5} and weight decay *w ∈* {10^−2^, 10^−3^}. Additionally, we tested stepwise, one-cycle and multi-cycle learning rate schedules and a light and a heavy augmenter. Initial values for the learning rate were set manually based on a learning rate sweep ^73^, though kept conservative to preserve the learned features from the pretraining. The hyperparameter search was implemented as a naive gridsearch over the total 24 different configurations (Supplementary Figure 9). We selected the model procuring the best AUC in the validation set to produce out-of-sample predictions for the outer hold-out fold. In the final evaluation of the models, we compiled predictions for all participants, for each using the model where they belonged to the hold-out test set. Our main method for measuring performance was the AUC, but we also report accuracy, which, due to our matching procedure, is equivalent to balanced accuracy.

### Relevance maps

We built a pipeline *LRP_dementia_* for generating relevance maps by implementing LRP (Bach et al., 2015) on top of the trained classifier. LRP is a technique for explaining single decisions made by the model, and thus, when running the pipeline on input a relevance map *R* is generated alongside the prediction 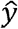. *R* is a three-dimensional volume, representing a visual explanation for 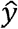, where each voxel *r_i,j,k_*, ∈ *R* has a spatial position *i,j,k* corresponding to the location of an input voxel *x_i,j,k_ ∈X*. Furthermore, the intensity of *r_i,j,k_* can be directly interpreted as how much voxel *x_i,j,k_* contributes to 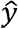, such that 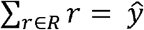. In the original LRP-formulation, relevance is propagated backwards between subsequent layers *Z_l_* and Z*_l+1_* according to the relative contribution of one artificial neuron *a_m_* ∈ *Z_l_* in the first layer on relevance in all artificial neurons *a_m_* ∈ Z*_l+1_* in the following layer,

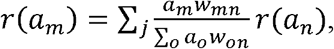

where *w*_mn_ denotes the weight between *a_m_* and *a_n_*. We controlled the influence of different aspects of the explanations using a composite LRP strategy ^44^, combining different formulations of the LRP-formula for the different layers in the model to enhance specific aspects of the relevance maps. Specifically, we employed a combination of alpha-beta and epsilon rules that have previously shown to produce meaningful results for dementia-classifiers ^41,42^, described in detail in the Supplementary Methods. The resulting relevance maps produced by the pipeline were full brain volumes with the same dimensionality as the MRI data (167×212×160 voxels) containing mostly (see below) positive relevance.

Notation-wise we generally consider the relevance map *R (X)* for an image *X* to be a function of the model *m_task_*, where *task* indicates which task the model was trained for, the LRP strategy LRP *_composite_* and the image *X*,

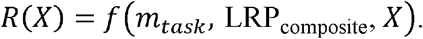

Because the composite LRP strategy described above is kept fixed in our pipeline, we contract this to

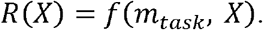

Furthermore, we let the model-specifier task annotate the map for a further simplification,

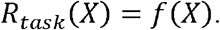

Thus, *LRP_task_* is used to annotate the full pipeline for a given task, while *R_task_* (*X*) denotes a single relevance map generated by this pipeline for image *X*. When the task is given by the context, we sometimes simplify this further to *R* (*X*), and when a general image is considered, we simply use *R* to denote its relevance map.

While we generally discuss our pipeline as a singular one, there were in reality five approximately equivalent pipelines (corresponding to the models trained for the five test folds), and which one is used depends on what image was used as input. Specifically, for each participant diagnosed with dementia, the pipeline is chosen where the participant was part of the hold-out test set while training the model, and both the relevance maps and the predictions are thus always out-of-sample. For participants used in the MCI analysis, which are all out-of-sample for all models, we created an ensemble by averaging the predictions and the voxel-wise relevance across all models

Before implementing the LRP procedure we made two slight modifications to the models to facilitate the backwards relevance propagation, both leaving the functional interface of the model unchanged. First, we removed the sigmoid activation in the final layer, so that the output of the model changed from a bounded continuous variable 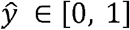 to an unbounded prediction 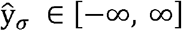. In this space a raw prediction of 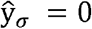is equivalent to a sigmoid-transformed prediction of 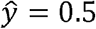, and thus 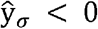means that the model predicts control status for the given participant, and oppositely 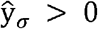implies that the model predicts a dementia diagnosis. Furthermore, this means that all positive relevance *r* ∈ *R, r >*0 can be interpreted as visual evidence in favour of a dementia diagnosis. Secondly, we modified the model by fusing all batch normalization layers with their preceding convolutional layers, adjusting their weights and biases to match the shift and scaling previously performed by the normalization layer ^74,75^.

After generation, the relevance maps are in the same stereotaxic space as their corresponding, linearly registered, input MRIs. To ensure intra-individual comparisons were done in the same space we non-linearly registered the maps to MNI152-space before subsequent statistical analyses were run. First, we registered the preprocessed MRIs used as inputs to the 1mm MNI152 template packaged with FSL using fnirt with splineorder=2. We then applied the transformation computed for *X* to *R* (*X*) using applywarp. We also restrained our relevance maps to contain strictly positive relevance, evidence in favour of a dementia prediction, by clipping them to a minimum value of 0. Furthermore, to remove edge-effects from our analyses, we enforce that there is no relevance in non-brain tissue by nullifying all relevance outside the brain:

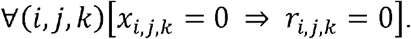

All visualized relevance maps are plotted after non-linear registration, overlayed on the MNI152-template. As the maps are three-dimensional, we generally plot a collection of distributed axial slices. The relevance is coloured by the nibabel v3.2.2 ^76^ cold_hot colourmap. Since the absolute relevance values vary between maps, all maps are normalized to the intensity range [0, 1] in the visualizations.

### Validating the relevance maps

Earlier studies have shown that interpretability techniques in general are prone to generate visual explanations that do not capture salient parts of the input ^51,52^. To investigate the extent of this for our pipeline *LRP_dementia_* we employed two analyses to assess the sanity of the relevance maps.

The first was an established task-specific technique comparing the relevance maps to existing knowledge of the pathology of dementia ^40^. The second was a purely quantitative analysis examining how important the regions found by the pipeline are for the dementia prediction 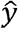. In both cases we contrasted the relevance maps generated from the main pipeline with three alternative pipelines representing variants of a null hypothesis, all expected to produce relevance maps with no significant association to dementia.

*LRP_random images_* represents the simplest alternative pipeline, and is built around the dementia-model, but with an additional preprocessing step scrambling the input,

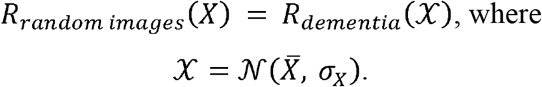

*LRP_random images_* is expected to generated relevance maps where the relevance is evenly distributed across the entire image. In the next pipeline *LRP_random weights_* we replaced the dementia-model with a model with random weights,

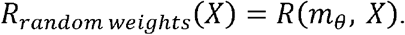

*m*_θ_ has not been trained for any task, and thus has random weights initialized by the default Keras “*Glorot Uniform*” weight-initializer. This pipeline is expected to produce relevance maps which correlate with the raw voxel intensities, e.g. high intensity in the input should entail more (absolute) relevance, thereby reflecting aspects of morphology. The final and most realistic alternative pipeline was *LRP*_sex_, where we replaced the dementia-model with a binary sex-classifier,

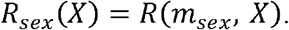

The sex-classifier was trained to differentiate males from females in one of the splits from the dementia-dataset, achieving an out-of-sample AUC of 0.956 and a balanced accuracy of 89.40%. We did not do any hyperparameter optimization for this model but used the best configuration from the dementia cross-validation in the same fold. The heatmaps from this pipeline should reflect regions where there is intra-individual variation in morphology, which are predictive of sex but with minimal association with dementia.

As a proxy for existing knowledge in the literature we performed an ALE meta-analysis using Sleuth v3.0.4 ^77^ and GingerALE v3.0.2 ^45^. We used Sleuth to search for relevant articles with the query

Imaging Modality is MRI AND

Context is disease AND

Diagnosis is Dementia OR Alzheimer’s Disease OR Lewy Body Dementia OR Frontotemporal Dementia OR Non-Aphasic Frontotemporal Dementia in the Voxel-based morphometry database, yielding 394 experiments from 124 articles. These experiments contained 3972 foci, 280 of which were outside the MNI152 mask, leaving 3692 to be loaded into GingerALE. Then the reference map *G*, with voxels *g _i.j,k_*, was generated by an ALE meta-analysis using the default parameters: Cluster-level FWE=0.01, Threshold Permutations=1000, P Value=0.001. The reference map is visualized in Supplementary Figure 4.

We performed four pairwise comparisons to estimate the amount of overlap between each of the pipelines and, *G*. For each pipeline the comparison was performed by computing an average map 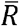, binarizing both it and *G*, and computing the Dice overlap between the two. The employed approach closely resembles the method of Wang et al. ^40^, but with multiple thresholds of binarization also for, and allowed us to plot similarity as a function of the threshold. The full details of the procedure is described in the Supplementary Methods. Additionally, to have a singular numerical basis for comparison, we computed the normalized cross-correlation ^46^ between the (non-binarized) average maps 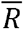 and the reference map *G*,

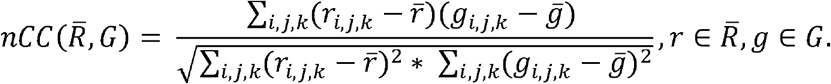

To facilitate an intuitive understanding of what parts of the brain the dementia-model is focusing on, we also performed a similar, region-wise comparison. This was done by extracting a subset of voxels from the average relevance map 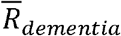,

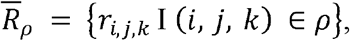

Where *ρ* is one of 69 regions defined in the Harvard-Oxford cortical and subcortical atlases ^78^. We did the same for *G* and let the mean activation per region for both constitute a tuple

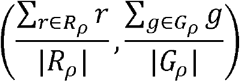

plotted Figure 2c. However, since it is non-trivial to determine which aggregation method corresponds to the most understandable and intuitive interpretation, we also created plots for tuples of sums,

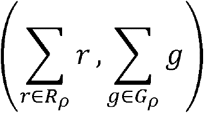

and maximum values

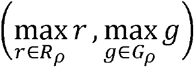

per region in Supplementary Figure 10.

To quantify the importance of the spatial locations captured by the various LRP pipelines for predicting dementia, we implemented a procedure for iteratively occluding parts of the image based on the relevance maps and observing how the prediction from the dementia model changed ^79^. Still using the true positives, for each pipeline *LRP_task_* for each MRI *X*_0_ we generated a baseline dementia-prediction 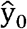and relevance map *R_task_*. Then we located the voxel with the highest amount of relevance in *R_task_* and replaced a 15×15×15 cube centred around the voxel with random uniform noise 𝒰(0,1), effectively concealing all information contained in this region. Next, we ran the modified image 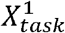 through the dementia-model to see how the prediction 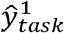 changed as a function of the occlusion. Note that injecting a box of random noise into the image is not trivially equivalent to removing information, however we specifically applied the same modification in the random box-augmentation during training and are thus hopeful that the model is invariant to the injection beyond the information removal. We iteratively applied this modify-and-predict procedure, also masking out the regions from the relevant maps between each iteration to minimize overlap of occlusion windows, for 20 iterations, producing a list of predictions 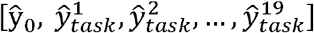 plotted for each pipeline in Figure 2d (averaged across all true positives). The rate of decline in these traces indicate the importance of the regions found in the respective relevance maps. We quantified the differences between the pipelines *LRP_task_* by calculating the area over the area over their perturbation curves ^79^,

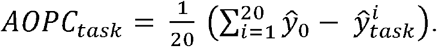

### Exploratory analyses in the MCI cohort

In the exploratory MCI analyses we used *LRP_dementia_* to generate predictions and relevance maps for participants from ADNI who were given an MCI diagnosis at inclusion. We first compiled the predictions and relevance maps (and the corresponding timestamps) for each participant at all timepoints into a single data structure we called a morphological record. We then tried to utilize this data structure to differentiate three groups: stable MCI patients (sMCI), progressive MCI patients (pMCI), and those who saw improvement in their cognition throughout the data collection phase. The remaining participants, e.g. those who either passed through all three diagnostic stages, or bounced between diagnoses, were excluded. Furthermore, we combined the stable and improving cohorts into a non-progressive group (nMCI) to facilitate binary group comparisons in the subsequent analyses.

For the first analysis comparing predictions in the two groups, due to variability in the total number and the frequency of visits between participants, we aimed to create a matched dataset based on visit history from the nMCI and pMCI cohorts to compare the predictions in the two groups with reference to a specific timepoint. We first started with all the progressive patients *p_p_* ∈ *pMCI* who got a diagnosis at timepoint *t_n+1_*, and, for each patient individually, compiled all previous visits *t_m_, m* ≤ *n* into a vector *h_p_* representing the time of the visits. The entries 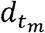 of the vector were the number of days until the diagnosis was given, *t_n+1_*− *t_m_*.For simplicity we also appended 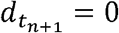 to *h*_p_, such that for a single patient

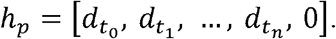

Then, for each of the non-progressive patients *p_p_* ∈ *nMCI* who didn’t have a time of diagnosis(e.g. *t_n+1_* is not given) we compiled a set *H_b_* of all possible history vectors *h_b_* by varying which visit was chosen as *t*_0_ and a terminal non-diagnosis timepoint *t_n+1_*. Next, we defined a cost-criterion for matching two histories (with an equal number of visits) as the sum of absolute pairwise differences between the vectors,

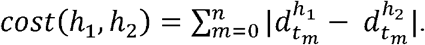

For each pair of progressive and non-progressive patients (*p_p_, p_n_*)this allowed us to calculate a best possible match, given that the stable patient had a total number of visits equal to or larger than the number of visits for the progressive patient:

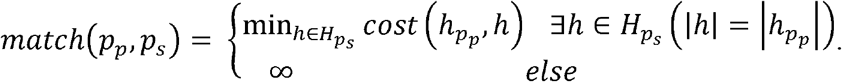

Finally, we compiled the cost of the optimal match from all pairs into a matrix and found the best complete matching by minimizing the total cost across this matrix using the Hungarian algorithm implemented in scipy v1.6.3 ^80^, such that each patient occurs in at most one pair.

We estimated differences in predictions 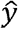 between the two groups using a linear mixed model. Specifically, we modelled 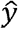 at all timepoints before the terminal timepoint *t_n+1_* as a function of age, sex (as controlling variables), years to diagnosis, categorical group membership (nMCI, pMCI), and an interaction between years to diagnosis and group. In addition, we had an independent intercept and slope per participant. The model was fit the formula API of statsmodels v0.13.2 ^81^ using the formula

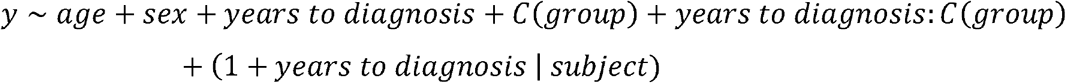

on the matched dataset. A full overview of coefficients and p-values can be found in Supplementary Table 4.

Due to the high dimensionality of the relevance maps, we decomposed them with a principal component analysis (PCA) before the final analyses. To fit the PCA we used the non-linearly registered relevance maps from a randomly selected timepoint for all MCI patients. Before fitting the model, all relevance maps were smoothed with a constant 3×3×3 blurring kernel using the convolution operation from Tensorflow 2.6 to strengthen the signal-to-noise ratio. The PCA was computed using scikit-learn v1.0.2 ^82^, retaining 64 components (out of 1137 maximally possible) in a component vector *c*=[*c*_0_,*c*_1_,…,*c*_63_].. An axial slice from each of the 64 components visualized in MNI152-space is shown in Supplementary Figure 6.

We fit Cox proportional hazard models using the component vectors as predictors to assess the association between the relevance maps and progression as a function of age. In addition to the components, representing the maps, we controlled for sex in the model. The p-values and coefficients can be found in Supplementary Table 5. To account for covariance between the components and the dementia-prediction 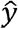 we ran an additional model where we divided the patients into ten strata based on 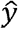. Both models were fit using lifelines v0.27.1 ^83^.

To further explore the prognostic efficacy of our pipeline we set up a predictive analysis for predicting progression at multiple, fixed timepoints a given number of months in the future. For each participant *p* with visits at timepoints *t^p^*, we denoted the last timepoint with an MCI diagnosis 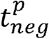 and the first timepoint with a dementia diagnosis (if present) 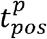. Using a fixed set of years into the future, γ ∈ {1,2,3,4,5}, we constructed a target variable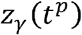 such that

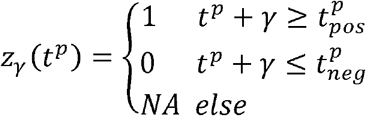

where the NAs allow for exclusion of all patients where the status at timepoint *t^p^+γ*is unknown. For each *γ* we constructed the target vector *z*_γ_ across all timepoints for all participants with *z*_γ=*NA*_ and split the constituent patients ρ into five folds stratified on *z*_γ_, sex and age, such that all timepoints from a participant resided in the same fold. Using these folds, we fit logistic regression models to predict *z*_γ_ with an *l*_1_-penalty in a nested cross-validation loop, allowing us to both tune the regularization parameter λ and have out-of-sample predictions for all participants. For eligible participants we used all timepoints for training the models, but during testing we sampled a random timepoint per participant to ensure independence between datapoints in the final evaluation. For each γ we fit three models: a baseline model

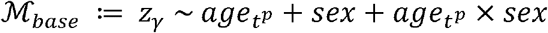

to assess the bias in the dataset with respect to age at the given timepoint *t^p^* and sex, a model using the prediction 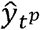 from the dementia classifier at *t^p^* as a predictor

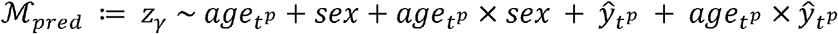

and a model including the relevance maps from *t^p^*, represented by the component vector 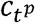,

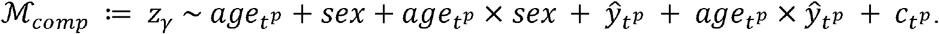

All models were fit and tuned using the LogisticRegressionCV interface of sklearn v1.0.2 ^82^. We compared models by measuring the mean AUC across the five folds (Supplementary Table 6).

To evaluate clinical applicability we also report accuracy, positive predictive value, sensitivity, and specificity (Table 2). To determine whether the more complex models represented significant improvements we employed a one-sided Wilcoxon signed-rank test from scipy v1.9.3 ^80^ to do pairwise comparisons between ℳ *_base_* and ℳ _*pred*_, and ℳ _*pred*_, and ℳ _*comp*_ across the five out-of-sample AUCs independently.

To assess whether the relevance maps were associated with specific cognitive functions we associated aspects of them with performance on various cognitive tests. We first extracted test results from seven neuropsychological batteries which spanned all ADNI waves and contained high-level summary scores from the ADNI website (Supplementary Table 7). We then manually extracted 17 summary scores spanning different, but overlapping, cognitive domains (Supplementary Table 8). The component vectors *c* were used as proxies for the relevance maps, where each *c_i_* represented a template for localization of pathology. We matched 2402 component vectors with test results from 733 MCI patients, forming a basis for the comparison. We then calculated the univariate association between cognitive performance according to each of the 17 with each of the dimensions *c_i_ ∈ c*, while including age and sex as covariates for correction. To isolate the effect of the localization we also corrected for dementia-prediction, 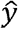. When a patient had multiple potential matches, a random timepoint was selected, and the final number of datapoints used in the analyses varied from 518 to 675. Correction for multiple testing was done with the Benjamini-Hochberg procedure. To ensure the associations were not confounded by collinearities between *c* and 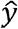, we also performed an equivalent analysis without correction to observe whether the sign of the coefficients changed.

## Data Availability

The raw data incorporated in this work were gathered from various resources. Material requests will need to be placed with individual principal investigators. A detailed overview of the independent datasets, and their origins, is provided in the supplementary information.

https://adni.loni.usc.edu/

http://www.aibl.csiro.au/

https://www.nitrc.org/projects/miriad/

https://www.oasis-brains.org/

## Author contributions

Conceptualization: EHL, TW, LTW, YW. Data curation: KP, EW, GS. Formal analysis: EHL. Funding acquisition: OAA, YW. Investigation: EHL, JMR, DVP, TK, AM, OAA, TW, LTW, YW. Methodology: EHL, EG, ND, TS, ØS, TW, LTW, YW. Project administration: GS, OAA, LTW, YW. Software: EHL. Supervision: TW, LTW, YW. Validation: EHL. Visualization: EHL. Writing – original draft: EHL, TW, LTW, YW. Writing – review & editing: KP, EG, ND, TS, JMR, DVP, ØS, TK, EW, AM, GS, OAA.

## Acknowledgements

This work was funded by the UiO:LifeScience Convergence Environment (project: 4MENT), the Research Council of Norway (302854), and the European Research Council under the European Union’s Horizon 2020 research and Innovation program (802998). The Southern and Eastern Norway Regional Health Authority supported the study through funding for KP but was not involved in conducting the study or in preparation of the manuscript. TW acknowledges funding from the German Research Foundation (DFG) Emmy Noether: 513851350. The work was performed on the Service for Sensitive Data (TSD) platform, owned by the University of Oslo, operated, and developed by the TSD service group at the University of Oslo IT-Department (USIT). We also acknowledge the computational resources provided by UNINETT Sigma2 - the National Infrastructure for High Performance Computing and Data Storage in Norway – with project no. (nn9769k/ns9769k).

## Data availability

The data used in this study were gathered from various sources, an overview including acknowledgements of their respective funding sources is provided in Supplementary Table 1. Among others, data used in the preparation of this article was obtained from the Alzheimer’s Disease Neuroimaging Initiative (ADNI, see adni.loni.usc.edu for further details), the Australian Imaging Biomarkers and Lifestyle flagship study of ageing (AIBL, www.aibl.csiro.au) the AddNeuroMed consortium, and MIRIAD (www.nitrc.org/projects/miriad). The investigators within these studies contributed to the design and implementation of the data collection process but did not participate in the analysis or writing of this report, and this publication is solely the responsibility of the authors.

## Code availability

The trained model and explainable pipeline and the underlying code will be made available at https://github.com/estenhl/pyment-public upon publication. Generic code for generating explanations for 3D CNNs is available at https://github.com/estenhl/keras-explainability.

