## Supplementary Methods, Figures and Tables for "Constructing personalized characterizations of structural brain aberrations in patients with dementia and mild cognitive impairment using explainable artificial intelligence"

Esten H. Leonardsen et al.

### Supplementary methods

#### LRP-strategy

To highlight specific facets of the relevance maps we employed a composite LRP strategy. For the prediction layer we retained the most salient explanations through an $LRP_\epsilon$-rule:

$$r_\epsilon(a_m) = \sum_n \left( \frac{a_m w_{mn}}{\epsilon + \sum_o a_o w_{on}} \right) r(a_n)$$

For the central convolutional layers, we upweighted positive relevance (e.g. features increasing the prediction, corresponding to evidence for a diagnosis) with

$$r_{\alpha\beta}(a_m) = \sum_n \left( \alpha \frac{(a_m w_{mn})^+}{\sum_o ((a_o w_{on})^+)} - \beta \frac{(a_m w_{mn})^-}{\sum_o ((a_o w_{on})^-)} \right) r(a_n)$$

where $(\cdot)^+$ and $(\cdot)^-$ denote positive and negative contributions respectively. For the input layer and the following convolutional layer we employed $LRP_\flat$ (also denoted as $LRP_{flat}$), to smooth finer details of the relevance maps:

$$r_\flat(a_m) = \sum_n \frac{1}{o} r(a_n)$$

where $o$ denotes the number of nodes connected to $r(a_n)$.

#### Comparison between relevance maps and GingerALE

For each pipeline, we first computed an average relevance map $\bar{R}$ across all true positives (e.g. dementia patients where the dementia-model correctly predicted a diagnosis, n=697), by computing their voxel-wise average relevance. Next, we binarized both the average map and the reference map by thresholding them at multiple percentiles $p \in [0, 100)$,

$$\bar{R}_p = \begin{cases} 1 & r_{i,j,k} > percentile(\bar{R}, p) \\ 0 & else \end{cases}$$

$$G_p = \begin{cases} 1 & g_{i,j,k} > percentile(G, p) \\ 0 & else \end{cases}$$

Then, for each percentile $p$ we calculate the Sørensen-Dice coefficient $SDC_p$ between the two,

$$SDC_p(\bar{R}_p, G_p) = \frac{\sum_{i,j,k} r_{i,j,k} g_{i,j,k}}{\sum_{i,j,k} r_{i,j,k} + \sum_{i,j,k} g_{i,j,k}}, r \in \bar{R}, g \in G_p.$$

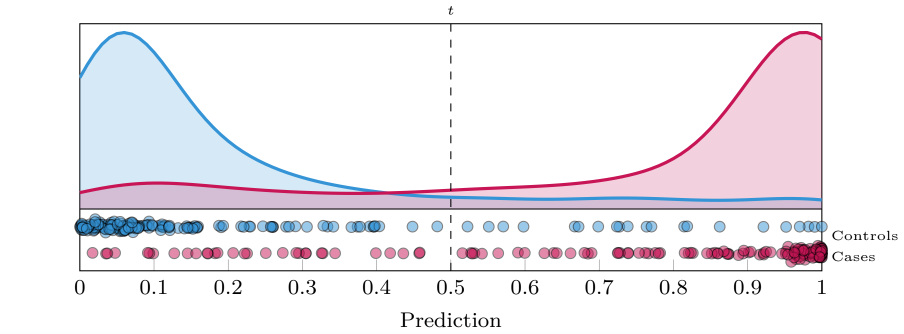

(a) Predictions

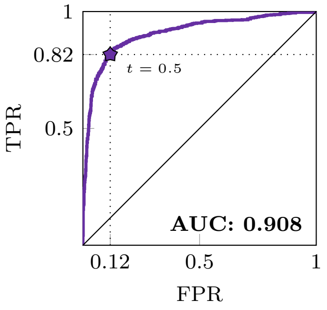

(b) ROC Curve

Predicted

| Observed | 0 | 1 |
|---|---|---|
| **0** | 754 | 100 |
| **1** | 157 | 697 |

**Accuracy: 84.95%**

(c) Confusion matrix

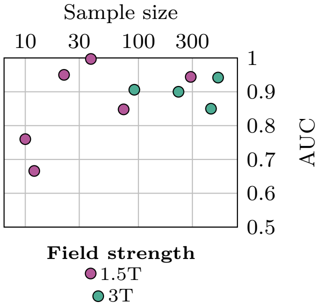

(d) AUC per site

Supplementary Figure 1: Predictive performance of the best performing dementia classifiers, combined across each out-of-sample test fold.

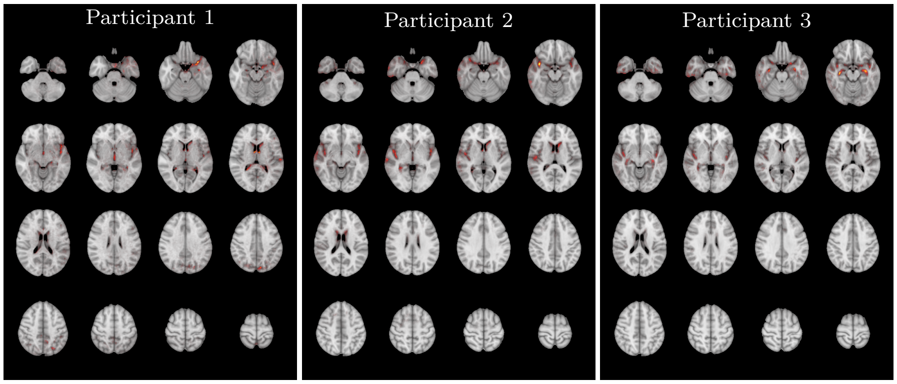

Supplementary Figure 2: Example relevance maps produced by the pipeline for three randomly selected dementia patients.

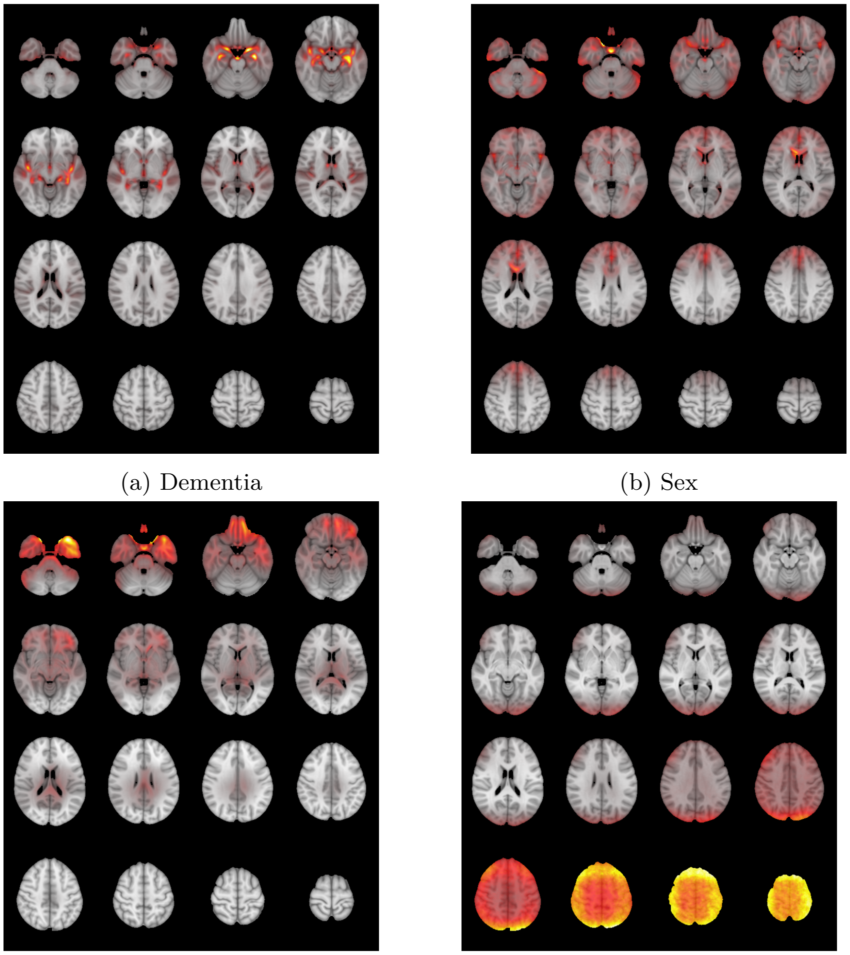

(a) Dementia

(b) Sex

(c) Random weights

(d) Randomized images

Supplementary Figure 3: Average voxel-wise relevance maps produced for the four pipelines across all correctly predicted dementia patients.

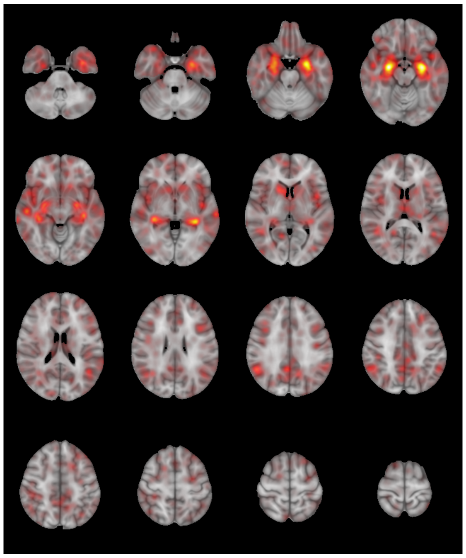

Supplementary Figure 4: Statistical map of dementia-related pathology generated by a GingerALE meta-analysis of 124 articles.

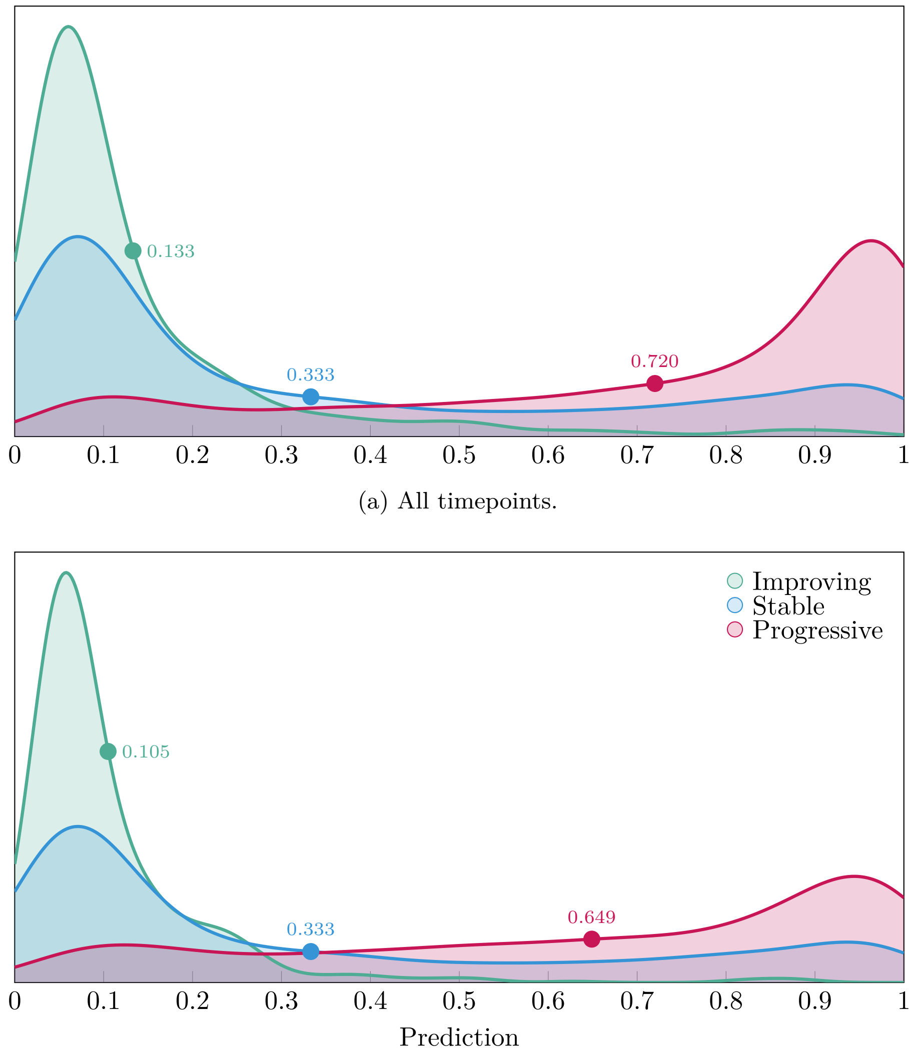

(a) All timepoints.

(b) Only timepoints when the patient got an MCI diagnosis.

Supplementary Figure 5: Distribution of predictions from the dementia classifier in the three MCI subsets. The marks indicate the mean prediction in each subset.

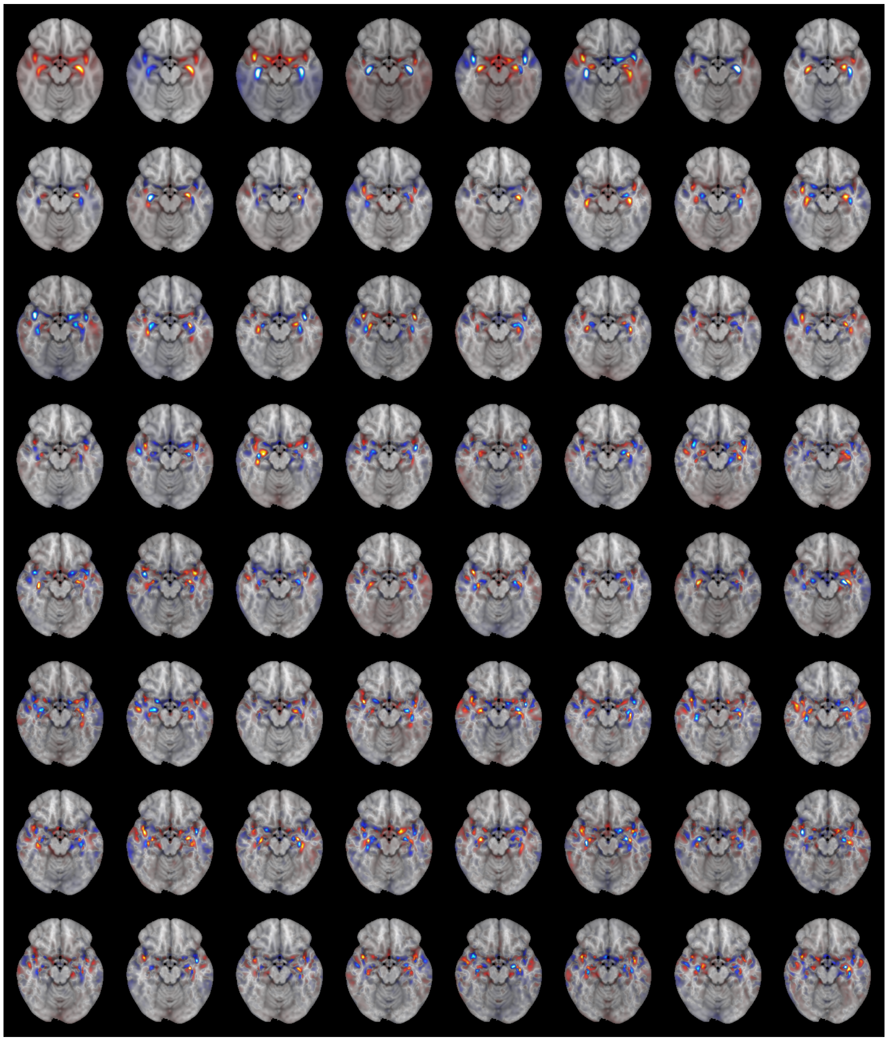

Supplementary Figure 6: A single axial slice from each of the 64 principal components plotted in brain space.

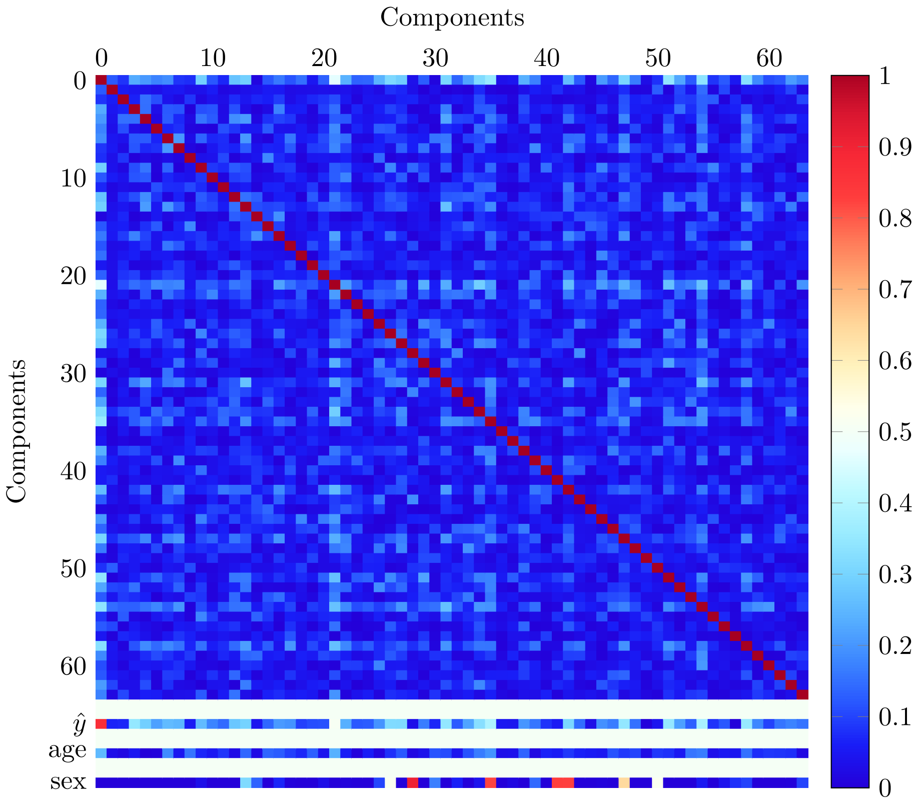

Supplementary Figure 7: Magnitudes of the correlations within the 64 principal components from the PCA, and between the components and prediction, age and sex. Correlations were computed in the dataset used for the exploratory MCI analyses.

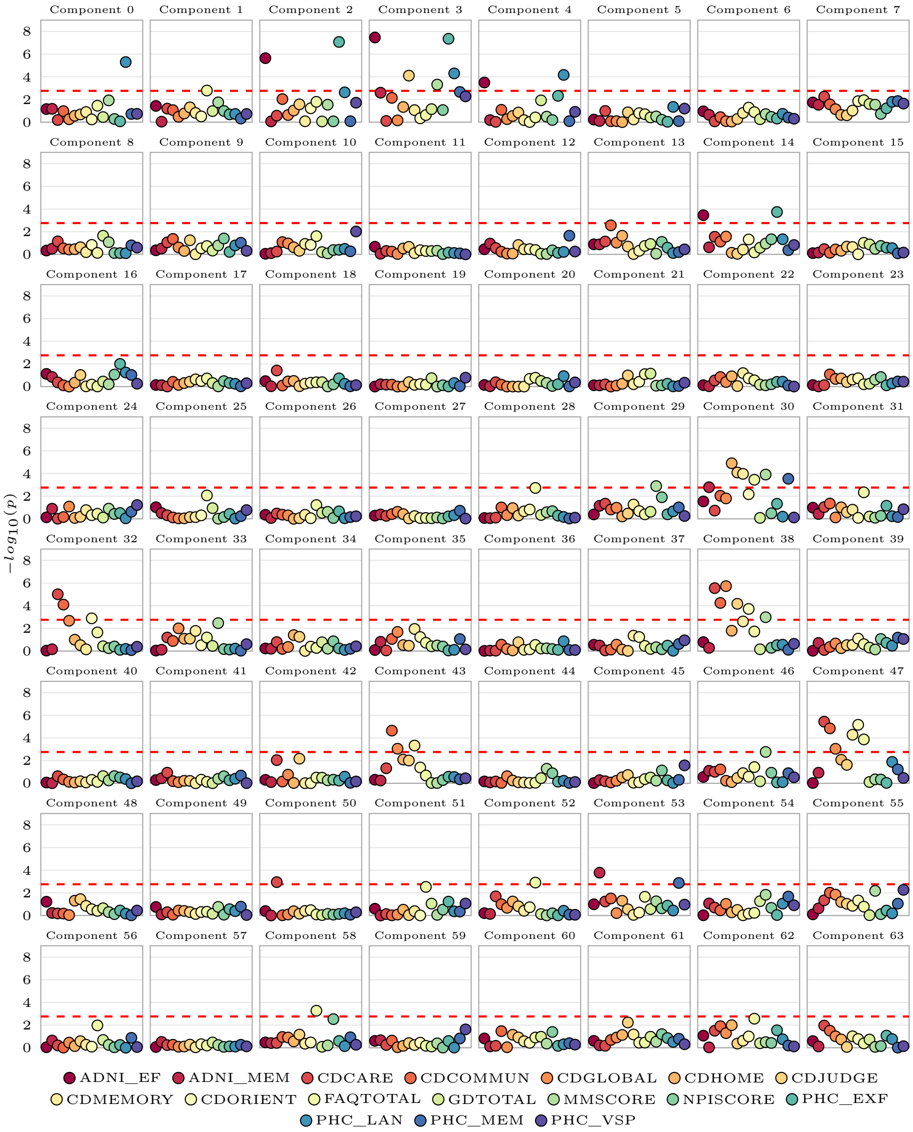

Supplementary Figure 8: P-values of the associations between the neuropsychological scores and each of the 64 components.

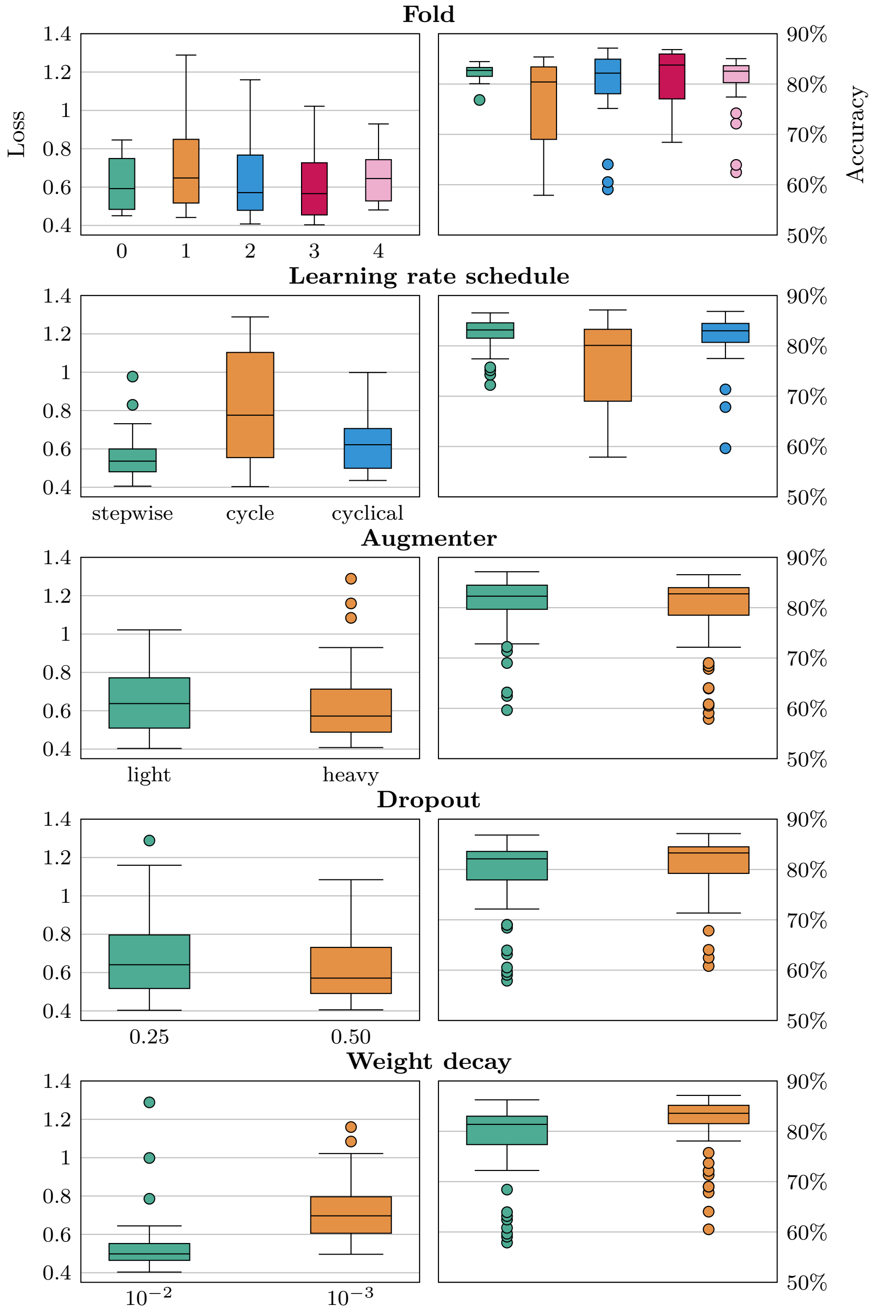

Supplementary Figure 9: Results achieved in the validation folds for the training runs of the dementia classifiers, grouped by each possible setting of each individual hyperparameter.

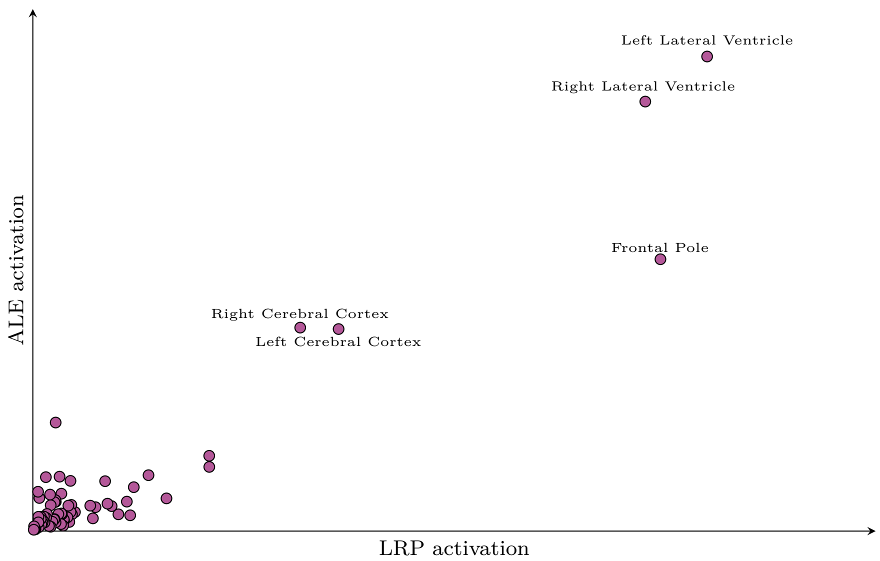

(a) Total activation per region

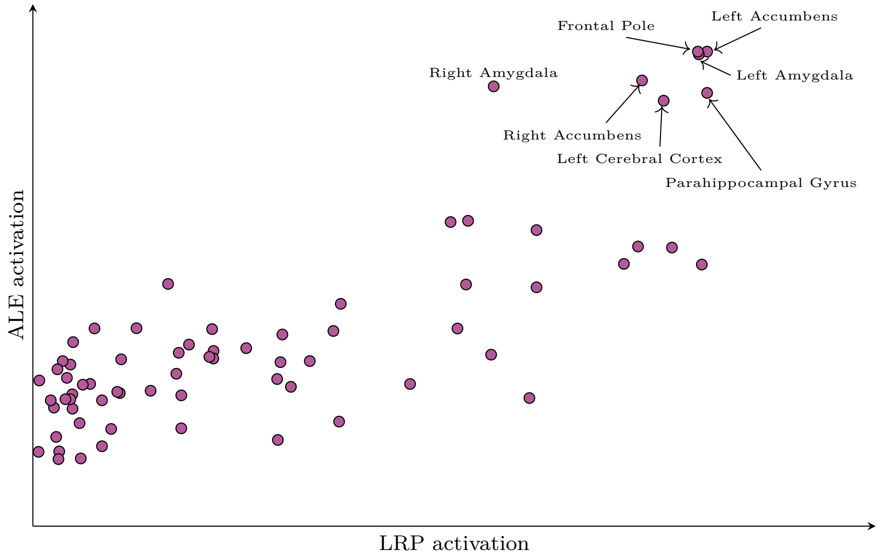

(b) Maximum activation per region

Supplementary Figure 10: Region-wise comparison of activation in the average map $\bar{R}_{dementia}$ produced by $LRP_{dementia}$ and $G$ produced by the GingerALE meta-analysis, using different aggregation methods.

| Cohort | Source | Comment | References |
|---|---|---|---|
| AddNeuroMed | Authors | AddNeuroMed consortium was led by Simon Lovestone, Bruno Vellas, Patrizia Mecocci, Magda Tsolaki, Iwona Kłoszewska, Hilkka Soininen. Their work was supported by InnoMed (Innovative Medicines in Europe), an integrated project funded by the European Union of the Sixth Framework program priority (FP6-2004- LIFESCIHEALTH-5) | [1, 2] |
| Alzheimer's Disease Neuroimaging Initiative | adni.loni.usc.edu | Data collection and sharing for this project was funded by the Alzheimer's Disease Neuroimaging Initiative (ADNI) (National Institutes of Health Grant U01 AG024904) and DOD ADNI (Department of Defense award number W81XWH-12-2-0012). ADNI is funded by the National Institute on Aging, the National Institute of Biomedical Imaging and Bioengineering, and through generous contributions from the following: AbbVie, Alzheimer's Association; Alzheimer's Drug Discovery Foundation; Araclon Biotech; BioClinica, Inc.; Biogen; Bristol-Myers Squibb Company; CereSpir, Inc.; Cogstate; Eisai Inc.; Elan Pharmaceuticals, Inc.; Eli Lilly and Company; EuroImmun; F. Hoffmann-La Roche Ltd and its affiliated company Genentech, Inc.; Fujirebio; GE Healthcare; IXICO Ltd.; Janssen Alzheimer Immunotherapy Research & Development, LLC.; Johnson & Johnson Pharmaceutical Research & Development LLC.; Lumosity; Lundbeck; Merck & Co., Inc.; Meso Scale Diagnostics, LLC.; NeuroRx Research; Neurotrack Technologies; Novartis Pharmaceuticals Corporation; Pfizer Inc.; Piramal Imaging; Servier; Takeda Pharmaceutical Company; and Transition Therapeutics. The Canadian Institutes of Health Research is providing funds to support ADNI clinical sites in Canada. Private sector contributions are facilitated by the Foundation for the National Institutes of Health (www.fnih.org). The grantee organization is the Northern California Institute for Research and Education, and the study is coordinated by the Alzheimer's Therapeutic Research Institute at the University of Southern California. ADNI data are disseminated by the Laboratory for Neuro Imaging at the University of Southern California. | [3, 4, 5] |
| Australian Imaging Biomarkers and Lifestyle flagship study of ageing | http://www.aibl.csiro.au/ | Data used in the preparation of this article was obtained from the Australian Imaging Biomarkers and Lifestyle flagship study of ageing (AIBL) funded by the Commonwealth Scientific and Industrial Research Organisation (CSIRO) which was made available at the ADNI database (www.loni.usc.edu/ADNI). The AIBL researchers contributed data but did not participate in analysis or writing of this report. AIBL researchers are listed at www.aibl.csiro.au | [6] |
| Demgen | Authors | Supported by the Norwegian National Advisory Unit on Aging and Health | [7, 8] |
| MIRIAD | https://www.nitrc.org/projects/miriad/ | Data used in the preparation of this article were obtained from the MIRIAD database (http://miriad.drc.ion.ucl.ac.uk). The MIRIAD investigators did not participate in analysis or writing of this report. The MIRIAD dataset is made available through the support of the UK Alzheimer's Society (Grant RF116). The original data collection was funded through an unrestricted educational grant from GlaxoSmithKline (Grant 6GKC). | [9] |
| OASIS3 | https://www.oasis-brains.org/ | Data were provided by OASIS-3: Longitudinal Multimodal Neuroimaging: Principal Investigators: T. Benzinger, D. Marcus, J. Morris; NIH P30 AG066444, P50 AG00561, P30 NS09857781, P01 AG026276, P01 AG003991, R01 AG043434, UL1 TR000448, R01 EB009352. AV-45 doses were provided by Avid Radiopharmaceuticals, a wholly owned subsidiary of Eli Lilly | [10] |
| StrokeMRI | Authors | Supported by the Research Council of Norway (249795, 248238), the South-Eastern Norway Regional Health Authority (2014097, 2015044, 2015073, 2016083), and the Norwegian ExtraFoundation for Health and Rehabilitation (2015/FO5146) | [11] |
| TOP | Authors | Supported by several grants from the Research Council of Norway, and the South-Eastern Norway Regional Health Authority | [12, 13, 14] |

Supplementary Table 1: An overview of the datasets used in the present study.

| **Dataset** | **Controls** | **Patients** |
|---|---|---|
| AddNeuroMed | MMSE $\geq$ 24 | MMSE < 19 |
| ADNI | DX = CN | DX = AD |
| AIBL | DX = DXNORM | DX $\in$ {DXAD, DXOTHDEM} |
| Demgen | - | DX $\in$ {AD, OtherDem, UnspecDem, VaD} |
| MIRIAD | Group = Control | Group = AD |
| OASIS3 | NORMCOG = 1 | NORMCOG = 0 & DEMENTED = 1 |
| StrokeMRI | Group = Control | - |
| TOP | diagnosis = CTRL | - |

Supplementary Table 2: Criteria for inclusion in the case-control groups. For replicability, variable and category names are kept as they were originally used in the originating dataset.

| | Validation | | Test | |
|---|---|---|---|---|
| **Fold** | **AUC** | **Accuracy** | **AUC** | **Accuracy** |
| 0 | 0.914 | 84.16% | 0.917 | 83.04 |
| 1 | 0.929 | 84.79% | 0.920 | 86.25 |
| 2 | 0.925 | 85.96% | 0.915 | 87.13 |
| 3 | 0.929 | 87.42% | 0.904 | 84.45 |
| 4 | 0.904 | 84.16% | 0.905 | 83.87 |
| | **0.918** | **85.30** | **0.908** | **84.95** |

Supplementary Table 3: Predictive performance of the dementia classifiers split up into individual folds. The fold number denotes which fold was used as the test fold for the given run.

| **Variable** | **Coefficient** | **p** |
|---|---|---|
| intercept | $-0.373$ | $1.94 \times 10^{-3}$ |
| sex[M] | $-0.084$ | $7.29 \times 10^{-4}$ |
| group[pMCI] | 0.473 | $6.05 \times 10^{-71}$ |
| age | 0.009 | $8.76 \times 10^{-10}$ |
| years_to_diagnosis | 0.013 | $3.92 \times 10^{-3}$ |
| years_to_diagnosis:group[pMCI] | 0.050 | $8.14 \times 10^{-17}$ |

Supplementary Table 4: Coefficients and p-values from the linear mixed model modelling dementia prediction $\hat{y}$ as a function of various variables, including membership in the progressive (pMCI) or non-progressive group.

| | Uncorrected | | Corrected for $\hat{y}$ | |
|---|---|---|---|---|
| **Component** | **p** | $\beta$ | **p** | $\beta$ |
| 0 | **$1.60 \times 10^{-66}$** | 0.683 | **0.003** | 0.18 |
| 1 | 0.061 | −0.043 | **0.001** | −0.07 |
| 2 | **$4.41 \times 10^{-26}$** | −0.248 | **$1.74 \times 10^{-20}$** | −0.22 |
| 3 | **$2.32 \times 10^{-20}$** | 0.222 | **$9.49 \times 10^{-10}$** | 0.16 |
| 4 | **$6.61 \times 10^{-14}$** | 0.173 | **$3.91 \times 10^{-09}$** | 0.14 |
| 5 | 0.117 | −0.032 | 0.525 | −0.01 |
| 6 | **$1.22 \times 10^{-04}$** | −0.083 | **$2.84 \times 10^{-05}$** | −0.09 |
| 7 | 0.11 | −0.036 | 0.037 | 0.05 |
| 8 | **$2.90 \times 10^{-04}$** | 0.076 | **$7.93 \times 10^{-04}$** | 0.07 |
| 9 | 0.164 | 0.028 | 0.766 | 0.01 |
| 10 | 0.024 | 0.051 | 0.049 | 0.05 |
| 11 | **$5.74 \times 10^{-05}$** | 0.082 | **$1.25 \times 10^{-04}$** | 0.08 |
| 12 | **$3.50 \times 10^{-05}$** | −0.09 | **$2.78 \times 10^{-06}$** | −0.1 |
| 13 | **0.003** | −0.063 | **$1.13 \times 10^{-05}$** | −0.1 |
| 14 | **$6.83 \times 10^{-05}$** | 0.088 | **0.003** | 0.07 |
| 15 | **$1.94 \times 10^{-04}$** | −0.081 | **$1.91 \times 10^{-06}$** | −0.1 |
| 16 | **0.001** | −0.071 | 0.016 | −0.05 |
| 17 | **$6.44 \times 10^{-05}$** | −0.087 | **0.001** | −0.07 |
| 18 | **0.002** | −0.064 | **$9.70 \times 10^{-07}$** | −0.1 |
| 19 | 0.316 | −0.022 | 0.152 | −0.03 |
| 20 | 0.492 | 0.016 | 0.949 | 0.0 |
| 21 | 0.484 | −0.018 | 0.374 | −0.02 |
| 22 | **0.008** | −0.067 | **$6.99 \times 10^{-05}$** | −0.1 |
| 23 | **0.018** | 0.049 | 0.068 | 0.04 |
| 24 | **$4.03 \times 10^{-06}$** | −0.106 | **$1.02 \times 10^{-04}$** | −0.09 |
| 25 | **0.017** | 0.057 | **$5.75 \times 10^{-04}$** | 0.08 |
| 26 | **$1.40 \times 10^{-07}$** | −0.11 | **$3.63 \times 10^{-07}$** | −0.11 |
| 27 | 0.368 | 0.019 | **0.006** | 0.06 |
| 28 | 0.546 | 0.013 | 0.229 | 0.03 |
| 29 | **$4.03 \times 10^{-04}$** | −0.075 | **$2.80 \times 10^{-04}$** | −0.08 |
| 30 | **$3.44 \times 10^{-08}$** | −0.104 | **$1.72 \times 10^{-15}$** | −0.16 |
| 31 | 0.859 | 0.004 | 0.617 | 0.01 |
| 32 | 0.061 | 0.04 | 0.029 | 0.05 |
| 33 | **0.003** | −0.063 | **$5.71 \times 10^{-04}$** | −0.08 |
| 34 | 0.059 | 0.04 | **0.001** | 0.07 |
| 35 | 0.316 | −0.024 | 0.792 | 0.01 |
| 36 | 0.233 | 0.023 | **0.007** | 0.05 |
| 37 | **$1.82 \times 10^{-05}$** | −0.09 | **$5.41 \times 10^{-06}$** | −0.1 |
| 38 | **$7.93 \times 10^{-04}$** | 0.075 | **0.001** | 0.07 |
| 39 | 0.267 | −0.022 | 0.07 | −0.04 |
| 40 | **0.01** | 0.052 | **$3.62 \times 10^{-05}$** | 0.08 |
| 41 | 0.159 | −0.034 | 0.08 | −0.04 |
| 42 | **$1.87 \times 10^{-07}$** | −0.116 | **$2.83 \times 10^{-10}$** | −0.14 |
| 43 | 0.044 | 0.042 | 0.104 | 0.03 |
| 44 | **0.009** | 0.058 | 0.1 | 0.04 |
| 45 | 0.157 | 0.032 | 0.287 | 0.02 |
| 46 | 0.81 | 0.005 | 0.399 | −0.02 |
| 47 | **$2.24 \times 10^{-05}$** | 0.087 | **0.001** | 0.07 |
| 48 | 0.067 | 0.034 | **0.009** | 0.05 |
| 49 | 0.159 | 0.028 | **0.009** | 0.05 |
| 50 | 0.969 | 0.001 | 0.196 | −0.02 |
| 51 | **0.004** | 0.065 | **0.01** | 0.06 |
| 52 | 0.691 | 0.008 | 0.424 | 0.02 |
| 53 | 0.805 | −0.005 | 0.941 | −0.0 |
| 54 | 0.204 | 0.032 | 0.071 | 0.05 |
| 55 | 0.082 | 0.033 | 0.121 | 0.03 |
| 56 | 0.207 | 0.028 | 0.104 | 0.04 |
| 57 | 0.147 | −0.03 | 0.052 | −0.04 |
| 58 | **$2.18 \times 10^{-08}$** | −0.11 | **$4.09 \times 10^{-09}$** | −0.12 |
| 59 | 0.05 | 0.035 | **$7.21 \times 10^{-04}$** | 0.06 |
| 60 | 0.412 | −0.017 | 0.173 | −0.03 |
| 61 | 0.554 | 0.011 | 0.555 | 0.01 |
| 62 | **$8.24 \times 10^{-04}$** | 0.067 | 0.023 | 0.05 |
| 63 | 0.336 | 0.019 | **0.006** | 0.05 |

Supplementary Table 5: Coefficient and p-value for each component in the two Partial Hazard analyses. In the corrected analyses (the two rightmost columns), the patients were stratified based on $\hat{y}$, to isolate the effect of the components.

| Model | AUC | Balanced accuracy | PPV | Sensitivity | Specificity |
|---|---|---|---|---|---|
| $\mathcal{M}_{base}$ | 0.506±0.039 | 49.85%±3.7% | 0.11±0.1 | 0.31±0.26 | 0.69±0.26 |
| $\mathcal{M}_{pred}$ | 0.666±0.083 | 61.49%±10.58% | 0.21±0.12 | 0.56±0.31 | 0.67±0.18 |
| $\mathcal{M}_{comp}$ | 0.743±0.063 | 70.1%±4.79% | 0.32±0.03 | 0.68±0.11 | 0.72±0.02 |

(a) One year

| Model | AUC | Balanced accuracy | PPV | Sensitivity | Specificity |
|---|---|---|---|---|---|
| $\mathcal{M}_{base}$ | 0.474±0.05 | 49.47%±5.34% | 0.33±0.05 | 0.47±0.06 | 0.52±0.07 |
| $\mathcal{M}_{pred}$ | 0.742±0.053 | 69.92%±5.24% | 0.52±0.07 | 0.75±0.09 | 0.65±0.08 |
| $\mathcal{M}_{comp}$ | 0.786±0.019 | 74.31%±0.99% | 0.61±0.04 | 0.72±0.04 | 0.76±0.06 |

(b) Two years

| Model | AUC | Balanced accuracy | PPV | Sensitivity | Specificity |
|---|---|---|---|---|---|
| $\mathcal{M}_{base}$ | 0.536±0.032 | 53.9%±3.78% | 0.32±0.26 | 0.35±0.29 | 0.73±0.23 |
| $\mathcal{M}_{pred}$ | 0.797±0.037 | 75.76%±4.16% | 0.75±0.04 | 0.76±0.09 | 0.76±0.06 |
| $\mathcal{M}_{comp}$ | 0.808±0.016 | 77.17%±2.29% | 0.79±0.04 | 0.72±0.03 | 0.82±0.05 |

(c) Three years

| Model | AUC | Balanced accuracy | PPV | Sensitivity | Specificity |
|---|---|---|---|---|---|
| $\mathcal{M}_{base}$ | 0.529±0.056 | 53.01%±3.74% | 0.38±0.32 | 0.35±0.29 | 0.71±0.24 |
| $\mathcal{M}_{pred}$ | 0.844±0.041 | 80.6%±4.09% | 0.84±0.04 | 0.82±0.06 | 0.79±0.04 |
| $\mathcal{M}_{comp}$ | 0.867±0.031 | 80.38%±3.26% | 0.87±0.02 | 0.75±0.08 | 0.85±0.02 |

(d) Four years

| Model | AUC | Balanced accuracy | PPV | Sensitivity | Specificity |
|---|---|---|---|---|---|
| $\mathcal{M}_{base}$ | 0.515±0.031 | 51.05%±2.09% | 0.14±0.28 | 0.09±0.18 | 0.93±0.14 |
| $\mathcal{M}_{pred}$ | 0.889±0.024 | 83.61%±1.81% | 0.91±0.02 | 0.83±0.04 | 0.84±0.03 |
| $\mathcal{M}_{comp}$ | 0.903±0.034 | 84.1%±2.52% | 0.92±0.02 | 0.82±0.03 | 0.86±0.04 |

(e) Five years

Supplementary Table 6: Predictive performance for each of the progression models at each of the five years following the MRI examination.

| Name | Included | Exclusion reason |
|---|---|---|
| Alzheimer's Disease Assessment Scale Sub-scores | No | Missing ADNI2, 3, GO |
| ADSP Phenotype Harmonization Consortium | Yes | |
| Alzheimer's Disease Assessment Scale | No | Missing description |
| Clinical Dementia Rating | Yes | |
| Cognitive Change Index | No | Missing ADNI1, GO |
| Cogstate Battery | No | Missing ADNI1, GO |
| Cogstate Brief Battery | No | Missing ADNI1, 2, GO |
| Digital Cognitive Biomarkers | No | Missing summary scores |
| Everyday Cognition | No | Missing ADNI1 |
| Financial Capacity Instrument | No | Missing ADNI1, 2, GO |
| Functional Activities Questionnaire | Yes | |
| Geriatric Depression Scale | Yes | |
| Item level data | No | Missing ADNI2, 3, GO |
| Mini-Mental State Examination | Yes | |
| Modified Haschinski Ischemia Scale | No | Non-congruent visit codes |
| Montreal Cognitive Assessment | No | Missing ADNI1 |
| Neuropsychiatric Inventory Questionnaire | Yes | |
| Neuropsychological Battery | No | Missing explanation |
| UW - Neuropsych Summary Scores | Yes | |

Supplementary Table 7: Neuropsychological assessments used as a basis for finding associations with the components from the relevance maps.

| Variable | Source | Description |
|---|---|---|
| PHC_MEM | ADSP Phenotype Harmonization Consortium | Harmonized composite memory score |
| PHC_EXF | ADSP Phenotype Harmonization Consortium | Harmonized composite executive function score |
| PHC_LAN | ADSP Phenotype Harmonization Consortium | Harmonized composite language score |
| PHC_VSP | ADSP Phenotypxe Harmonization Consortium | Harmonized composite visuospatial score |
| CDRMEMORY | Clinical Dementia Rating | Clinical evaluation of impairment related to memory |
| CDRORIENT | Clinical Dementia Rating | Clinical evaluation of impairment related to orientation |
| CDRJUDGE | Clinical Dementia Rating | Clinical evaluation of impairment related to judgement and problem solving |
| CDRCOMMUN | Clinical Dementia Rating | Clinical evaluation of impairment related to community affairs |
| CDRHOME | Clinical Dementia Rating | Clinical evaluation of impairment related to home and hobbies |
| CDRCARE | Clinical Dementia Rating | Clinical evaluation of impairment related to personal care |
| CDRGLOBAL | Clinical Dementia Rating | Total clinical dementia rating |
| MMSCORE | Mini-Mental State Examination | Overall measure of cognitive impairment |
| FAQTOTAL | Functional Activities Questionnaire | Measures instrumental activities from daily life |
| GDTOTAL | Geriatric Depression Scale | Screening for depression in elderly adults |
| NPISCORE | Neuropsychiatric Inventory Questionnaire | Informant-based assessment of Neuropsychiatric symptoms |
| ADNI_EF | UW - Neuropsych Summary Scores | Composite score for executive functioning |
| ADNI_MEM | UW - Neuropsych Summary Scores | Composite score for memory |

Supplementary Table 8: Test scores which were correlated against the components from the relevance maps.

| **Hyperparameter** | **Values** |
|---|---|
| Dropout | {0.25, 0.5} |
| Weight decay | {1e-2, 1e-3} |
| Augmenter | {Light, Heavy} |
| Learning rate | {Stepwise, Cyclical, One-Cycle} |

Supplementary Table 9: Hyperparameters that were tuned when training the dementia classifiers.

[6] Kathryn A. Ellis et al. "The Australian Imaging, Biomarkers and Lifestyle (AIBL) study of aging: methodology and baseline characteristics of 1112 individuals recruited for a longitudinal study of Alzheimer's disease". In:

*International Psychogeriatrics* 21.4 (Aug. 2009), pp. 672–687. ISSN: 1041-6102. DOI: 10.1017/S1041610209009405.
